# Severe and common mental disorders and risk of hospital admissions for Ambulatory Care Sensitive Conditions (ACSCs): prospective cohort study using UK Biobank

**DOI:** 10.1101/2022.01.05.22268793

**Authors:** Claire L. Niedzwiedz, María José Aragón, Josefien J. F. Breedvelt, Daniel J Smith, Stephanie L. Prady, Rowena Jacobs

## Abstract

**Background:** People with mental disorders have an excess chronic disease burden. One mechanism to potentially reduce the public health and economic costs of mental disorders is to reduce preventable hospital admissions. Ambulatory care sensitive conditions (ACSCs) are a defined set of chronic and acute illnesses not considered to require hospital treatment if patients receive adequate primary healthcare. We examined the relationship between both severe and common mental disorders and risk of emergency hospital admissions for ACSCs and factors associated with increased risk.

**Methods:** Baseline data from England (N=445,814) were taken from UK Biobank, which recruited participants aged 37-73 years during 2006 to 2010, and were linked to hospital admission records up to 31^st^ December 2019. Participants were grouped into those who had a history of either schizophrenia, bipolar disorder, depression or anxiety, or no record of mental disorder. Cox proportional hazard models (for the first admission) and Prentice, Williams and Peterson Total Time models (PWP-TT, which account for all admissions) were used to assess the risk (using hazard ratios (HR)) of hospitalisation for ACSCs among those with mental disorders compared to those without, adjusting for factors in different domains, including sociodemographic (e.g. age, sex, ethnicity), socioeconomic (e.g. deprivation, education level), health and biomarkers (e.g. multimorbidity, inflammatory markers), health-related behaviours (e.g. smoking, alcohol consumption), social isolation (e.g. social participation, social contact) and psychological (e.g. depressive symptoms, loneliness).

**Results:** People with schizophrenia had the highest risk of hospital admission for ACSCs compared to those with no mental disorder (HR=4.40, 95% CI: 4.04 - 4.80). People with bipolar disorder (HR=2.48, 95% CI: 2.28 – 2.69) and depression or anxiety (HR=1.76, 95% CI: 1.73 – 1.80) also had higher risk. Associations were more conservative when accounting for all admissions. Although adjusting for a range of factors attenuated the observed associations, they still persisted, with socioeconomic and health-related variables contributing most.

**Conclusions:** People with severe mental disorders had highest risk of preventable hospital admissions, with the risk also elevated amongst those with depression and anxiety. Ensuring people with mental disorders receive adequate ambulatory care is essential to reduce the large health inequalities experienced by these groups.

## 1 Background

People with mental disorders have double the risk of mortality compared to the general population, with a decade of years of potential life lost.(1) The burden of mortality is highest among people with severe mental disorders (SMI; including schizophrenia, bipolar disorder and psychotic conditions), but is also elevated among those with common mental disorders (CMD), such as depression and anxiety.(2) Several studies have demonstrated that this excess mortality is mostly attributable to a higher burden of non-communicable diseases including cardiovascular disease, smoking-related lung disease and type II diabetes.(3-5) Potential explanations for this include poorer quality of health and social care, lower adherence to treatment for physical health conditions, side effects of psychotropic medications, unhealthier behaviours such as smoking, alcohol consumption and physical inactivity, as well as underlying social inequalities.(5, 6)

Hospitalisations for chronic illness represent a major proportion of overall healthcare spending.(7) Therefore, preventing hospital admissions is likely to yield economic benefits, as well as reducing the overall burden on the health service. Ambulatory Care Sensitive Conditions (ACSCs) are health conditions which are not considered to require inpatient treatment with appropriate management via primary care intervention.(8) In England, ACSCs represent a sixth of emergency admissions, with an annual cost of £1.42 billion to the NHS.(9) They therefore represent a key target for reduction, especially given the increasing trend over recent years.(10) ACSCs can be grouped into either acute (e.g. dehydration or gastroenteritis, where more severe progression can be prevented via early intervention), chronic (e.g. asthma, where effective care can reduce exacerbation of disease symptoms) or preventable (via vaccines and other interventions e.g. influenza or pneumonia).(9) Elevated levels of hospital admissions for ACSCs can be an indicator of poor continuity of care between primary and secondary care.(11)

Few studies have investigated hospital admissions for ACSCs among people with mental disorders, especially in the UK. Previous research from Denmark and Taiwan has demonstrated people with SMI have higher risk of ACSC admissions, compared to those without.(6, 12) A study based in New York state, limited by its cross-sectional design and restricted geographic coverage, found that people with mental disorders were two times more likely to be admitted to hospital due to an ACSC compared to those without a mental disorder.(13) This was similar to a study based on the population of Western Australia, which used linked data from 1990 to 2006 and found that mental health patients were two times more likely to experience potentially preventable hospital admissions.(14) Limitations of previous research include a lack of comparison between different mental disorders, with studies often either grouping all conditions together, or focusing on one specific condition only.(12, 15) Research has also been limited by the sole use of electronic health records,(6, 14, 15) which often do not contain sufficient data to investigate a range of potential confounding and mediating factors, such as income and social support. Most previous studies have also not taken into account the full burden of hospital admissions over time, often being restricted to the first admission or rehospitalisation within a certain time period.

### 1.1 Aims & Objectives

We aimed to examine the risk of emergency hospital admissions for ACSCs among people with a range of mental disorders and examine factors that potentially explain any excess risk. We analysed data from UK Biobank, a highly phenotyped data source that links self-report, hospitalisation, mortality and biological data, using standard as well as multiple-failure survival analysis techniques. The objectives were to:

- Examine risk of hospital admissions for ACSCs among individuals with and without severe and common mental disorders (SCMD; i.e. schizophrenia, bipolar disorder, depression and anxiety).
- Explore the factors (sociodemographic, socioeconomic, health and biomarkers, health-related behaviours, social isolation and psychological factors) that help to explain any increased risk.

## 2 Methods

### 2.1 Data

Secondary data were taken from UK Biobank (https://www.ukbiobank.ac.uk/), which achieved a 5.5% response rate.(16, 17) Over 502 000 community-dwelling individuals aged 37 to 73 years were recruited to UK Biobank during 2006 to 2010. Participants attended one of 22 assessment centres across England, Scotland and Wales. For this study we limited the sample to those attending assessment centres in England. Baseline assessments were linked to Hospital Episode Statistics (HES) for England and death records provided by NHS Digital (both up to 31^st^ December 2019). UK Biobank received ethical approval from the NHS National Research Ethics Service North West (21/NW/0157). All participants gave written informed consent before enrolment in the study, which was conducted in accordance with the principles of the Declaration of Helsinki. We excluded participants who requested their data be withdrawn from UK Biobank (updated on 09 August 2021).

### 2.2 Cohort definition

Individuals with a severe or common mental disorder (SCMD) were identified via linked clinical records or self-report using the ‘first occurrence’ data field (1712). UK Biobank provides the date on which a diagnosis was recorded for the first time and the source (e.g. primary care or hospital admission). Data from 45% of the UK Biobank cohort is linked to primary care records,(18) which use Read codes to record clinical events and these are converted to the corresponding International Classification of Diseases-10 (ICD-10) code by the UK Biobank team. For each diagnosis group of interest (bipolar disorder (ICD-10 codes F30; F31), schizophrenia and other psychotic disorders (F20-F29), depression (F32-39) and anxiety and related disorders (F40-F48)) the earliest date on which a diagnosis was recorded was identified.

At the baseline assessment centre individuals could also self-report (data field 20002) a lifetime diagnosis of ‘schizophrenia’, ‘mania/bipolar disorder/manic depression’, ‘depression’ or anxiety and related disorders: ‘anxiety/panic attacks’, ‘nervous breakdown’, ‘post-traumatic stress disorder’, ‘obsessive compulsive disorder’ or ‘stress’. A subset of UK Biobank participants (those recruited in 2009-2010) also completed detailed questionnaires about lifetime depressive and mania symptoms at the baseline assessment, from which probable cases of major depression and bipolar disorder have been derived by clinicians.(19) For solely self-reported records the date of diagnosis was recorded as the date the individual joined UK Biobank. If a person had more than one SCMD diagnosis, we ranked them in the following order: (1) schizophrenia, (2) bipolar, (3) anxiety or depression, and classified the patient according to the highest ranked. Participants who had an ICD-10 code under Chapter V (mental and behavioural disorders (e.g. eating disorders)) not covered by the mental disorder categories above, were excluded from the sample (n=31,923) and all those with no recorded mental or behavioural disorder were grouped into the control group. For the analysis we grouped those with anxiety or depression (common mental disorders) together due to their significant co-morbidity.(20)

### 2.3 Outcome

The primary outcome of interest was emergency hospital admissions for an Ambulatory Care Sensitive Condition (ACSC). ACSCs were defined according to the <u>NHS England criteria</u> which includes 19 conditions divided into acute, chronic and vaccine-preventable.(21) Acute conditions included cellulitis, dehydration, dental conditions, ear, nose and throat infections, gangrene, gastroenteritis, nutritional deficiency, pelvic inflammatory disease, ulcers, and urinary conditions. Chronic conditions included angina, asthma, chronic obstructive pulmonary disease, congestive heart failure, diabetes, epilepsy, hypertension, and anaemia. Vaccine preventable conditions included influenza, pneumonia, tuberculosis, hepatitis B, or other.

Hospital admissions for ACSCs were extracted from the Hospital Episode Statistics (HES) for England supplied by NHS Digital via UK Biobank.(22) HES reports data as episodes (period of care under a consultant) and there can be more than one episode during a hospitalisation. Each episode can record multiple diagnoses (via ICD-10 codes), which can be used to identify ACSCs. An ACSC admission was defined as an ACSC condition recorded in the first episode. Since we were interested in emergency ACSC admissions, we excluded elective and maternity admissions.

In order to identify ACSC admissions, we grouped together consecutive episodes of the same patient. UK Biobank does not report hospital codes, so these continuous periods in hospital can include transfers between different hospitals. To construct these “continuous inpatient spells (CIPS)” we used information about the source and method of admission and the discharge destination, together with the start and end dates of the episodes to make sure the episodes were in the correct order. Additional file Table S1 contains detail on the data exclusions in HES to identify ACSC admissions.

### 2.4 Covariates

We included a range of potential variables which may influence the association between SCMD and ACSC admissions, grouped into sociodemographic, socioeconomic, health and biomarkers, health-related behaviours, social isolation and psychological factors. All data for the covariates were collected at the baseline assessment centre

Sociodemographic factors included: age (years); sex (male, female), ethnicity (white British, white Irish, other white background, south Asian, black, mixed or other), urban/rural residence (based on home postcode population density) and assessment centre attended. Socioeconomic factors included education level (1: university or college degree, 2: A-levels or equivalent, 3: O-levels, General Certificate of Secondary Education (GCSE), vocational Certificate of Secondary Education (CSE) or equivalent, 4: other (e.g. National Vocational Qualifications or other professional qualifications), or 5: none of the above); deprivation at the output area level (assessed using the Townsend index (23), converted to a Z score where higher levels reflect higher levels of deprivation); employment status (paid employment or self-employment, retired, looking after home and/or family, unable to work because of sickness or disability, unemployment or other); housing tenure (owner-occupier or renter/other); household income (before tax, self-reported): <£18,000; £18,000-£30,999; £31,000-£51,999; £52,000-£100,000; or >£100,000.

Health measures included multimorbidity (number of self-reported chronic physical health conditions, top-coded at 4 or more, based on a previously published approach (24), excluding mental health conditions); body mass index (BMI) category (underweight, normal weight, overweight, obese). We included three biomarkers indicative of inflammation (C-reactive Protein (CRP), logged), metabolic (waist circumference) and cardiovascular function (pulse rate). Indicators of health behaviours included smoking (never, previous, current) and alcohol consumption (daily or almost daily, 3-4 times a week, once or twice a week, 1-3 times per month, special occasions, former drinker or never). Physical activity (walking, moderate and vigorous) in a typical week was also recorded using self-reported items from the International Physical Activity Questionnaire short form(25), from which a single measure of total physical activity in metabolic equivalent of task (MET) hours per week was derived; this was converted into quintiles.(26)

A number of measures were used to capture social isolation: living arrangements (with spouse/partner; with other people; live alone); social contact (visit friends/family less than weekly versus at least once a week); and social participation (one or more activity (e.g. sports club) at least once a week versus no activities at least once a week). Finally, psychological factors included loneliness (whether participants often feel lonely – yes or no), current depressive symptoms (measured using an adapted Patient Health Questionnaire (PHQ-4)(27) and sleeplessness (never/rarely, sometimes, usually).

### 2.5 Statistical analysis

First, descriptive statistics for the sample were calculated, including the number of hospital admissions by SCMD diagnosis. We then ran several survival models to assess the relationship between SCMD (schizophrenia, bipolar disorder, depression or anxiety, and those with no disorder as the referent group) and ACSC admissions in the following order:

1. unadjusted;
2. model 1 plus age, sex, ethnicity, urban/rural, assessment centre location (sociodemographic factors);
3. model 2 plus education, deprivation, employment status, housing tenure and income (socioeconomic factors);
4. model 3 plus multimorbidity, BMI, pulse rate, waist circumference and CRP (health and biomarker variables);
5. model 3 plus smoking, alcohol consumption, physical activity (health-related behaviours);
6. model 3 plus living arrangements, social participation and social contact (social isolation factors);
7. model 3 plus depressive symptoms, sleeplessness and loneliness (psychological factors);
8. all variables.

The observation period for each person started on the date of the initial baseline UK Biobank assessment centre attendance or when they were diagnosed with a SCMD, if the diagnosis was later than the assessment. During the observation period, a patient could have none, one or more than one hospital admission. Models were censored at the earliest date of ACSC admission, date of death or the end of follow-up on 31^st^ December 2019. We use two model specifications, one modelling the time to first admission (Cox proportional hazard model) and one that considered all admissions (Prentice, Williams and Peterson Total Time (PWP-TT) model).(28, 29) The time to first admission model does not use all data (it ignores second and later admissions) and can, therefore, show associations between risk factors and admissions that do not hold once all admissions are considered.(28) The PWP-TT analyses ordered multiple events via stratification, based on the prior number of events during the follow-up period.(29, 30) It therefore takes into account that having a prior admission affects the risk of future admission and that the effect of covariates may differ in subsequent events.(29) Further details on these models and how they can be implemented in Stata software can be found elsewhere.(28) Participants with missing data for any variable were excluded from the analysis (models therefore contain a different number of individuals). The extent of missing data varied from 1.5% in model 2 and 35.5% in model 8. Violations of the proportional hazard assumption were examined graphically by plotting scaled Schoenfeld residuals. Statistical analysis was performed using Stata/MP 17.

## 3 Results

### 3.1 Description of sample

Our sample comprised of 413,891 participants (Figure 1), who attended an assessment centre in England and had either no prior psychiatric disorder diagnosis (N=319,365), a previous diagnosis of schizophrenia (N=1,884), bipolar disorder (N=2,978) or anxiety/depression (N=89,664). Most participants with a SCMD experienced a hospital admission during the 13-year follow-up period, over half had emergency admissions and 10,832 participants had emergency ACSC admissions, with 7,218 experiencing just one admission and 504 experiencing more than five (Additional file Table S2). Table 1 shows the descriptive statistics (derived from the model containing all covariates, excluding missing data) by SCMD diagnosis. Across all diagnosis groups, individuals with SCMD were more likely to live in deprived areas, less likely to be in paid employment, more likely to experience poorer overall health, have adverse health behaviours and be socially isolated, compared to those without.

**Table 1:**
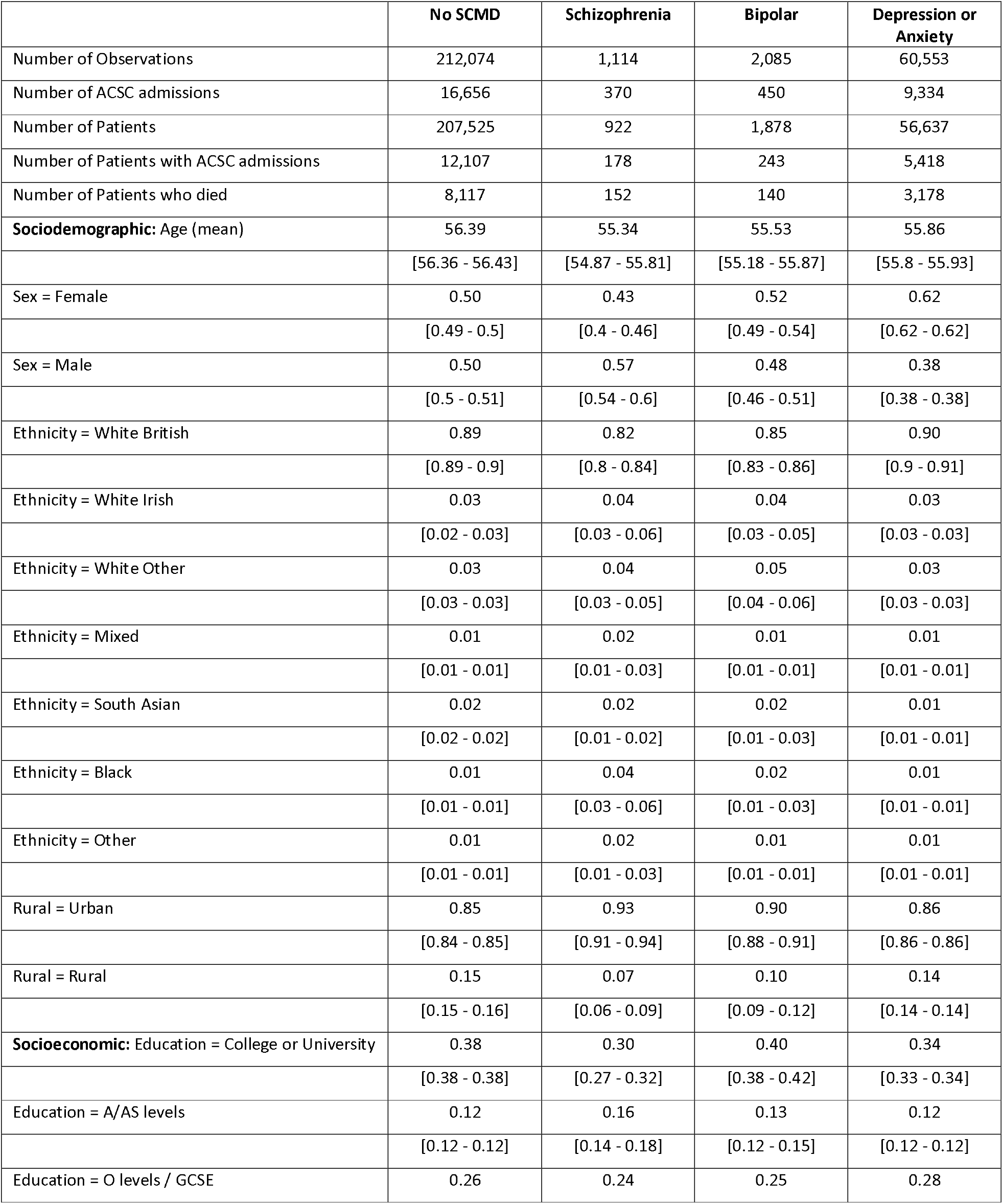
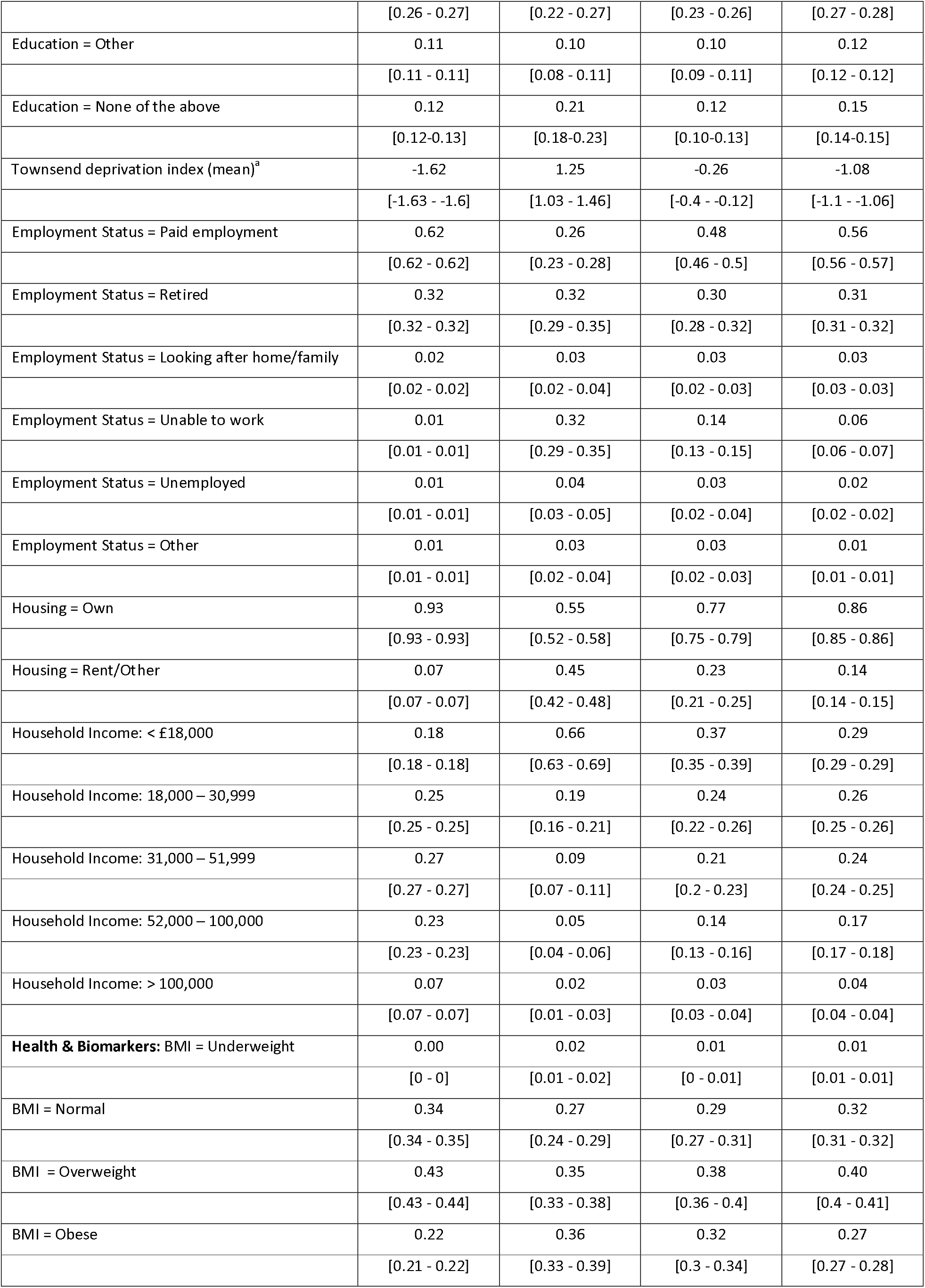
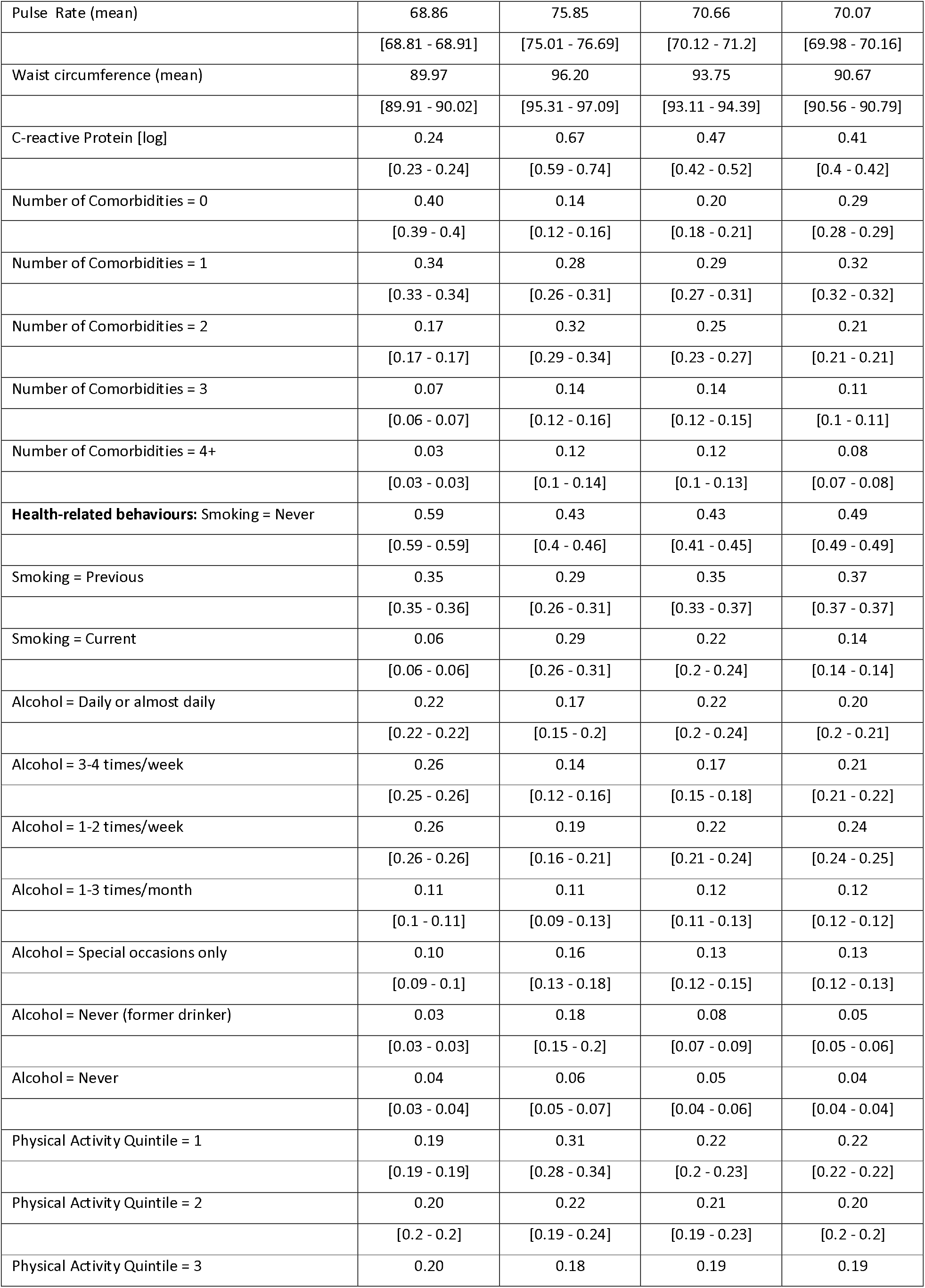
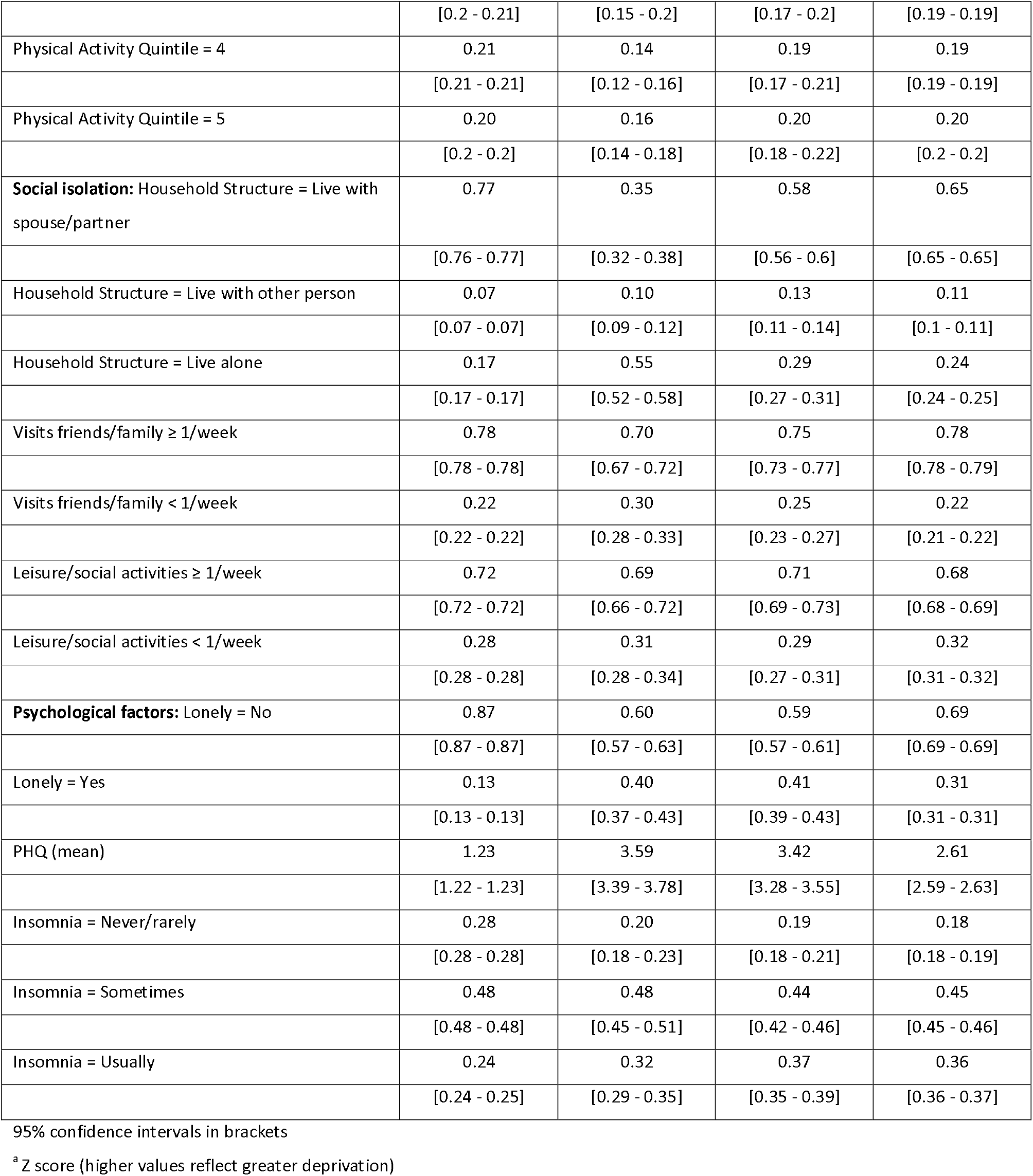
Descriptive statistics for the sample (proportions and 95% confidence intervals in brackets unless specified)

|  | No SCMD | Schizophrenia | Bipolar | Depression or Anxiety |
| --- | --- | --- | --- | --- |
| Number of Observations | 212,074 | 1,114 | 2,085 | 60,553 |
| Number of ACSC admissions | 16,656 | 370 | 450 | 9,334 |
| Number of Patients | 207,525 | 922 | 1,878 | 56,637 |
| Number of Patients with ACSC admissions | 12,107 | 178 | 243 | 5,418 |
| Number of Patients who died | 8,117 | 152 | 140 | 3,178 |
| **Sociodemographic:** Age (mean) | 56.39 | 55.34 | 55.53 | 55.86 |
|  | [56.36 - 56.43] | [54.87 - 55.81] | [55.18 - 55.87] | [55.8 - 55.93] |
| Sex = Female | 0.50 | 0.43 | 0.52 | 0.62 |
|  | [0.49 - 0.5] | [0.4 - 0.46] | [0.49 - 0.54] | [0.62 - 0.62] |
| Sex = Male | 0.50 | 0.57 | 0.48 | 0.38 |
|  | [0.5 - 0.51] | [0.54 - 0.6] | [0.46 - 0.51] | [0.38 - 0.38] |
| Ethnicity = White British | 0.89 | 0.82 | 0.85 | 0.90 |
|  | [0.89 - 0.9] | [0.8 - 0.84] | [0.83 - 0.86] | [0.9 - 0.91] |
| Ethnicity = White Irish | 0.03 | 0.04 | 0.04 | 0.03 |
|  | [0.02 - 0.03] | [0.03 - 0.06] | [0.03 - 0.05] | [0.03 - 0.03] |
| Ethnicity = White Other | 0.03 | 0.04 | 0.05 | 0.03 |
|  | [0.03 - 0.03] | [0.03 - 0.05] | [0.04 - 0.06] | [0.03 - 0.03] |
| Ethnicity = Mixed | 0.01 | 0.02 | 0.01 | 0.01 |
|  | [0.01 - 0.01] | [0.01 - 0.03] | [0.01 - 0.01] | [0.01 - 0.01] |
| Ethnicity = South Asian | 0.02 | 0.02 | 0.02 | 0.01 |
|  | [0.02 - 0.02] | [0.01 - 0.02] | [0.01 - 0.03] | [0.01 - 0.01] |
| Ethnicity = Black | 0.01 | 0.04 | 0.02 | 0.01 |
|  | [0.01 - 0.01] | [0.03 - 0.06] | [0.01 - 0.03] | [0.01 - 0.01] |
| Ethnicity = Other | 0.01 | 0.02 | 0.01 | 0.01 |
|  | [0.01 - 0.01] | [0.01 - 0.03] | [0.01 - 0.01] | [0.01 - 0.01] |
| Rural = Urban | 0.85 | 0.93 | 0.90 | 0.86 |
|  | [0.84 - 0.85] | [0.91 - 0.94] | [0.88 - 0.91] | [0.86 - 0.86] |
| Rural = Rural | 0.15 | 0.07 | 0.10 | 0.14 |
|  | [0.15 - 0.16] | [0.06 - 0.09] | [0.09 - 0.12] | [0.14 - 0.14] |
| **Socioeconomic:** Education = College or University | 0.38 | 0.30 | 0.40 | 0.34 |
|  | [0.38 - 0.38] | [0.27 - 0.32] | [0.38 - 0.42] | [0.33 - 0.34] |
| Education = A/AS levels | 0.12 | 0.16 | 0.13 | 0.12 |
|  | [0.12 - 0.12] | [0.14 - 0.18] | [0.12 - 0.15] | [0.12 - 0.12] |
| Education = O levels / GCSE | 0.26 | 0.24 | 0.25 | 0.28 |
|  | [0.26 - 0.27] | [0.22 - 0.27] | [0.23 - 0.26] | [0.27 - 0.28] |
| Education = Other | 0.11 | 0.10 | 0.10 | 0.12 |
|  | [0.11 - 0.11] | [0.08 - 0.11] | [0.09 - 0.11] | [0.12 - 0.12] |
| Education = None of the above | 0.12 | 0.21 | 0.12 | 0.15 |
|  | [0.12-0.13] | [0.18-0.23] | [0.10-0.13] | [0.14-0.15] |
| Townsend deprivation index (mean)<sup>a</sup> | -1.62 | 1.25 | -0.26 | -1.08 |
|  | [-1.63 - -1.6] | [1.03 - 1.46] | [-0.4 - -0.12] | [-1.1 - -1.06] |
| Employment Status = Paid employment | 0.62 | 0.26 | 0.48 | 0.56 |
|  | [0.62 - 0.62] | [0.23 - 0.28] | [0.46 - 0.5] | [0.56 - 0.57] |
| Employment Status = Retired | 0.32 | 0.32 | 0.30 | 0.31 |
|  | [0.32 - 0.32] | [0.29 - 0.35] | [0.28 - 0.32] | [0.31 - 0.32] |
| Employment Status = Looking after home/family | 0.02 | 0.03 | 0.03 | 0.03 |
|  | [0.02 - 0.02] | [0.02 - 0.04] | [0.02 - 0.03] | [0.03 - 0.03] |
| Employment Status = Unable to work | 0.01 | 0.32 | 0.14 | 0.06 |
|  | [0.01 - 0.01] | [0.29 - 0.35] | [0.13 - 0.15] | [0.06 - 0.07] |
| Employment Status = Unemployed | 0.01 | 0.04 | 0.03 | 0.02 |
|  | [0.01 - 0.01] | [0.03 - 0.05] | [0.02 - 0.04] | [0.02 - 0.02] |
| Employment Status = Other | 0.01 | 0.03 | 0.03 | 0.01 |
|  | [0.01 - 0.01] | [0.02 - 0.04] | [0.02 - 0.03] | [0.01 - 0.01] |
| Housing = Own | 0.93 | 0.55 | 0.77 | 0.86 |
|  | [0.93 - 0.93] | [0.52 - 0.58] | [0.75 - 0.79] | [0.85 - 0.86] |
| Housing = Rent/Other | 0.07 | 0.45 | 0.23 | 0.14 |
|  | [0.07 - 0.07] | [0.42 - 0.48] | [0.21 - 0.25] | [0.14 - 0.15] |
| Household Income: < £18,000 | 0.18 | 0.66 | 0.37 | 0.29 |
|  | [0.18 - 0.18] | [0.63 - 0.69] | [0.35 - 0.39] | [0.29 - 0.29] |
| Household Income: 18,000 – 30,999 | 0.25 | 0.19 | 0.24 | 0.26 |
|  | [0.25 - 0.25] | [0.16 - 0.21] | [0.22 - 0.26] | [0.25 - 0.26] |
| Household Income: 31,000 – 51,999 | 0.27 | 0.09 | 0.21 | 0.24 |
|  | [0.27 - 0.27] | [0.07 - 0.11] | [0.2 - 0.23] | [0.24 - 0.25] |
| Household Income: 52,000 – 100,000 | 0.23 | 0.05 | 0.14 | 0.17 |
|  | [0.23 - 0.23] | [0.04 - 0.06] | [0.13 - 0.16] | [0.17 - 0.18] |
| Household Income: > 100,000 | 0.07 | 0.02 | 0.03 | 0.04 |
|  | [0.07 - 0.07] | [0.01 - 0.03] | [0.03 - 0.04] | [0.04 - 0.04] |
| **Health & Biomarkers: BMI = Underweight** | 0.00 | 0.02 | 0.01 | 0.01 |
|  | [0 - 0] | [0.01 - 0.02] | [0 - 0.01] | [0.01 - 0.01] |
| BMI = Normal | 0.34 | 0.27 | 0.29 | 0.32 |
|  | [0.34 - 0.35] | [0.24 - 0.29] | [0.27 - 0.31] | [0.31 - 0.32] |
| BMI = Overweight | 0.43 | 0.35 | 0.38 | 0.40 |
|  | [0.43 - 0.44] | [0.33 - 0.38] | [0.36 - 0.4] | [0.4 - 0.41] |
| BMI = Obese | 0.22 | 0.36 | 0.32 | 0.27 |
|  | [0.21 - 0.22] | [0.33 - 0.39] | [0.3 - 0.34] | [0.27 - 0.28] |
| Pulse Rate (mean) | 68.86 | 75.85 | 70.66 | 70.07 |
|  | [68.81 - 68.91] | [75.01 - 76.69] | [70.12 - 71.2] | [69.98 - 70.16] |
| Waist circumference (mean) | 89.97 | 96.20 | 93.75 | 90.67 |
|  | [89.91 - 90.02] | [95.31 - 97.09] | [93.11 - 94.39] | [90.56 - 90.79] |
| C-reactive Protein [log] | 0.24 | 0.67 | 0.47 | 0.41 |
|  | [0.23 - 0.24] | [0.59 - 0.74] | [0.42 - 0.52] | [0.4 - 0.42] |
| Number of Comorbidities = 0 | 0.40 | 0.14 | 0.20 | 0.29 |
|  | [0.39 - 0.4] | [0.12 - 0.16] | [0.18 - 0.21] | [0.28 - 0.29] |
| Number of Comorbidities = 1 | 0.34 | 0.28 | 0.29 | 0.32 |
|  | [0.33 - 0.34] | [0.26 - 0.31] | [0.27 - 0.31] | [0.32 - 0.32] |
| Number of Comorbidities = 2 | 0.17 | 0.32 | 0.25 | 0.21 |
|  | [0.17 - 0.17] | [0.29 - 0.34] | [0.23 - 0.27] | [0.21 - 0.21] |
| Number of Comorbidities = 3 | 0.07 | 0.14 | 0.14 | 0.11 |
|  | [0.06 - 0.07] | [0.12 - 0.16] | [0.12 - 0.15] | [0.1 - 0.11] |
| Number of Comorbidities = 4+ | 0.03 | 0.12 | 0.12 | 0.08 |
|  | [0.03 - 0.03] | [0.1 - 0.14] | [0.1 - 0.13] | [0.07 - 0.08] |
| **Health-related behaviours: Smoking = Never** | 0.59 | 0.43 | 0.43 | 0.49 |
|  | [0.59 - 0.59] | [0.4 - 0.46] | [0.41 - 0.45] | [0.49 - 0.49] |
| Smoking = Previous | 0.35 | 0.29 | 0.35 | 0.37 |
|  | [0.35 - 0.36] | [0.26 - 0.31] | [0.33 - 0.37] | [0.37 - 0.37] |
| Smoking = Current | 0.06 | 0.29 | 0.22 | 0.14 |
|  | [0.06 - 0.06] | [0.26 - 0.31] | [0.2 - 0.24] | [0.14 - 0.14] |
| Alcohol = Daily or almost daily | 0.22 | 0.17 | 0.22 | 0.20 |
|  | [0.22 - 0.22] | [0.15 - 0.2] | [0.2 - 0.24] | [0.2 - 0.21] |
| Alcohol = 3-4 times/week | 0.26 | 0.14 | 0.17 | 0.21 |
|  | [0.25 - 0.26] | [0.12 - 0.16] | [0.15 - 0.18] | [0.21 - 0.22] |
| Alcohol = 1-2 times/week | 0.26 | 0.19 | 0.22 | 0.24 |
|  | [0.26 - 0.26] | [0.16 - 0.21] | [0.21 - 0.24] | [0.24 - 0.25] |
| Alcohol = 1-3 times/month | 0.11 | 0.11 | 0.12 | 0.12 |
|  | [0.1 - 0.11] | [0.09 - 0.13] | [0.11 - 0.13] | [0.12 - 0.12] |
| Alcohol = Special occasions only | 0.10 | 0.16 | 0.13 | 0.13 |
|  | [0.09 - 0.1] | [0.13 - 0.18] | [0.12 - 0.15] | [0.12 - 0.13] |
| Alcohol = Never (former drinker) | 0.03 | 0.18 | 0.08 | 0.05 |
|  | [0.03 - 0.03] | [0.15 - 0.2] | [0.07 - 0.09] | [0.05 - 0.06] |
| Alcohol = Never | 0.04 | 0.06 | 0.05 | 0.04 |
|  | [0.03 - 0.04] | [0.05 - 0.07] | [0.04 - 0.06] | [0.04 - 0.04] |
| Physical Activity Quintile = 1 | 0.19 | 0.31 | 0.22 | 0.22 |
|  | [0.19 - 0.19] | [0.28 - 0.34] | [0.2 - 0.23] | [0.22 - 0.22] |
| Physical Activity Quintile = 2 | 0.20 | 0.22 | 0.21 | 0.20 |
|  | [0.2 - 0.2] | [0.19 - 0.24] | [0.19 - 0.23] | [0.2 - 0.2] |
| Physical Activity Quintile = 3 | 0.20 | 0.18 | 0.19 | 0.19 |
|  | [0.2 - 0.21] | [0.15 - 0.2] | [0.17 - 0.2] | [0.19 - 0.19] |
| Physical Activity Quintile = 4 | 0.21 | 0.14 | 0.19 | 0.19 |
|  | [0.21 - 0.21] | [0.12 - 0.16] | [0.17 - 0.21] | [0.19 - 0.19] |
| Physical Activity Quintile = 5 | 0.20 | 0.16 | 0.20 | 0.20 |
|  | [0.2 - 0.2] | [0.14 - 0.18] | [0.18 - 0.22] | [0.2 - 0.2] |
| **Social isolation:** Household Structure = Live with spouse/partner | 0.77 | 0.35 | 0.58 | 0.65 |
|  | [0.76 - 0.77] | [0.32 - 0.38] | [0.56 - 0.6] | [0.65 - 0.65] |
| Household Structure = Live with other person | 0.07 | 0.10 | 0.13 | 0.11 |
|  | [0.07 - 0.07] | [0.09 - 0.12] | [0.11 - 0.14] | [0.1 - 0.11] |
| Household Structure = Live alone | 0.17 | 0.55 | 0.29 | 0.24 |
|  | [0.17 - 0.17] | [0.52 - 0.58] | [0.27 - 0.31] | [0.24 - 0.25] |
| Visits friends/family ≥ 1/week | 0.78 | 0.70 | 0.75 | 0.78 |
|  | [0.78 - 0.78] | [0.67 - 0.72] | [0.73 - 0.77] | [0.78 - 0.79] |
| Visits friends/family < 1/week | 0.22 | 0.30 | 0.25 | 0.22 |
|  | [0.22 - 0.22] | [0.28 - 0.33] | [0.23 - 0.27] | [0.21 - 0.22] |
| Leisure/social activities ≥ 1/week | 0.72 | 0.69 | 0.71 | 0.68 |
|  | [0.72 - 0.72] | [0.66 - 0.72] | [0.69 - 0.73] | [0.68 - 0.69] |
| Leisure/social activities < 1/week | 0.28 | 0.31 | 0.29 | 0.32 |
|  | [0.28 - 0.28] | [0.28 - 0.34] | [0.27 - 0.31] | [0.31 - 0.32] |
| **Psychological factors:** Lonely = No | 0.87 | 0.60 | 0.59 | 0.69 |
|  | [0.87 - 0.87] | [0.57 - 0.63] | [0.57 - 0.61] | [0.69 - 0.69] |
| Lonely = Yes | 0.13 | 0.40 | 0.41 | 0.31 |
|  | [0.13 - 0.13] | [0.37 - 0.43] | [0.39 - 0.43] | [0.31 - 0.31] |
| PHQ (mean) | 1.23 | 3.59 | 3.42 | 2.61 |
|  | [1.22 - 1.23] | [3.39 - 3.78] | [3.28 - 3.55] | [2.59 - 2.63] |
| Insomnia = Never/rarely | 0.28 | 0.20 | 0.19 | 0.18 |
|  | [0.28 - 0.28] | [0.18 - 0.23] | [0.18 - 0.21] | [0.18 - 0.19] |
| Insomnia = Sometimes | 0.48 | 0.48 | 0.44 | 0.45 |
|  | [0.48 - 0.48] | [0.45 - 0.51] | [0.42 - 0.46] | [0.45 - 0.46] |
| Insomnia = Usually | 0.24 | 0.32 | 0.37 | 0.36 |
|  | [0.24 - 0.25] | [0.29 - 0.35] | [0.35 - 0.39] | [0.36 - 0.37] |

95% confidence intervals in brackets
<sup>a</sup> Z score (higher values reflect greater deprivation)

**Figure 1:**
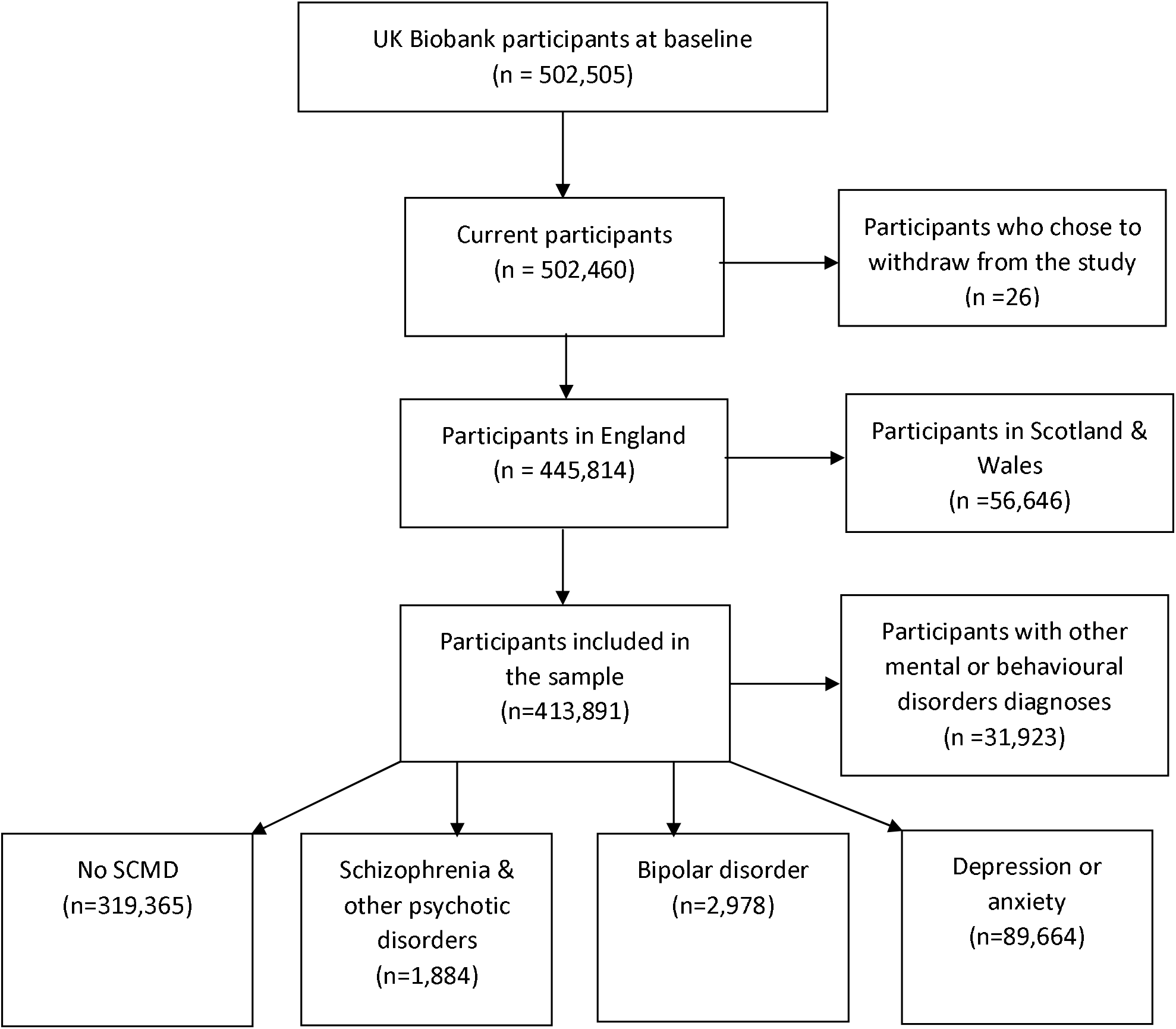
STROBE flowchart of study participants

### 3.2 Risk of ACSC hospital admissions

When looking at the first admission only in unadjusted models, people with schizophrenia had the highest risk of ACSC admission compared to those with no mental disorder (HR=4.40, 95% CI: 4.04 - 4.80) (Table 2). People with bipolar disorder (HR=2.48, 95% CI: 2.28 – 2.69) and depression or anxiety (HR=1.76, 95% CI: 1.73 – 1.80) also had heightened risk. When taking multiple admissions into account (Table 3) the associations were weaker but still elevated; schizophrenia (HR=2.29, 95% CI: 2.08 – 2.52), bipolar disorder (HR=1.92, 95% CI: 1.78 – 2.08), depression or anxiety (HR=1.57, 95% CI: 1.53 – 1.60).

**Table 2.** Results from Cox Proportional Hazard models for the association between severe and common mental disorders and risk of hospital admission for Ambulatory Care Sensitive Conditions (first admission per person only)

|  | **M1** | **M2** | **M3** | **M4** | **M5** | **M6** | **M7** | **M8** |
| --- | --- | --- | --- | --- | --- | --- | --- | --- |
|  | **HR**<br>**[95% CI]** | **HR**<br>**[95% CI]** | **HR**<br>**[95% CI]** | **HR**<br>**[95% CI]** | **HR**<br>**[95% CI]** | **HR**<br>**[95% CI]** | **HR**<br>**[95% CI]** | **HR**<br>**[95% CI]** |
| **Ref: No SCMD diagnosis** |  |  |  |  |  |  |  |  |
| Schizophrenia | 4.40 | 4.86 | 2.65 | 2.31 | 2.47 | 2.65 | 2.56 | 2.15 |
|  | [4.04-4.80] | [4.44-5.32] | [2.37-2.97] | [2.05-2.61] | [2.19-2.79] | [2.36-2.97] | [2.26-2.89] | [1.87-2.48] |
| Bipolar | 2.48 | 2.81 | 2.12 | 1.72 | 1.95 | 2.10 | 1.95 | 1.56 |
|  | [2.28-2.69] | [2.58-3.06] | [1.92-2.33] | [1.55-1.90] | [1.76-2.16] | [1.90-2.31] | [1.76-2.16] | [1.39-1.76] |
| Anxiety or Depression | 1.76 | 1.98 | 1.72 | 1.53 | 1.63 | 1.71 | 1.59 | 1.43 |
|  | [1.73-1.80] | [1.93-2.02] | [1.68-1.77] | [1.49-1.57] | [1.59-1.67] | [1.67-1.75] | [1.55-1.64] | [1.39-1.47] |
| **Adjustments** |  |  |  |  |  |  |  |  |
| Sociodemographic | No | Yes | Yes | Yes | Yes | Yes | Yes | Yes |
| Socioeconomic | No | No | Yes | Yes | Yes | Yes | Yes | Yes |
| Health & biomarkers | No | No | No | Yes | No | No | No | Yes |
| Health-related behaviours | No | No | No | No | Yes | No | No | Yes |
| Social isolation | No | No | No | No | No | Yes | No | Yes |
| Psychological | No | No | No | No | No | No | Yes | Yes |
| **Number of Observations** | 413,203 | 407,028 | 342,143 | 313,770 | 319,173 | 340,337 | 309,100 | 266,616 |
| **Number of Patients** | 413,203 | 407,028 | 342,143 | 313,770 | 319,173 | 340,337 | 309,100 | 266,616 |
| **Number of Failures** | 44,802 | 44,164 | 35,412 | 31,984 | 31,713 | 35,177 | 31,343 | 25,818 |
| **Log Likelihood** | -569,729 | -554,545 | -437,069 | -389,027 | -388,894 | -433,910 | -383,573 | -309,995 |

**Table 3:** Results from Prentice, Williams and Peterson Total Time (PWP-TT) models for the association between severe and common mental disorders and risk of hospital admissions for Ambulatory Care Sensitive Conditions (all admissions)

|  | **M1** | **M2** | **M3** | **M4** | **M5** | **M6** | **M7** | **M8** |
| --- | --- | --- | --- | --- | --- | --- | --- | --- |
|  | **HR**<br>**[95% CI]** | **HR**<br>**[95% CI]** | **HR**<br>**[95% CI]** | **HR**<br>**[95% CI]** | **HR**<br>**[95% CI]** | **HR**<br>**[95% CI]** | **HR**<br>**[95% CI]** | **HR**<br>**[95% CI]** |
| **Ref: No MD diagnosis** |  |  |  |  |  |  |  |  |
| Schizophrenia | 2.29 | 2.64 | 2.09 | 1.96 | 1.92 | 2.10 | 2.11 | 1.86 |
|  | [2.08-2.52] | [2.38-2.93] | [1.84-2.38] | [1.70-2.25] | [1.66-2.23] | [1.85-2.39] | [1.83-2.42] | [1.57-2.20] |
| Bipolar | 1.92 | 2.13 | 1.79 | 1.58 | 1.84 | 1.78 | 1.73 | 1.62 |
|  | [1.77-2.08] | [1.95-2.33] | [1.62-1.98] | [1.42-1.76] | [1.68-2.03] | [1.61-1.97] | [1.55-1.92] | [1.46-1.80] |
| Anxiety or Depression | 1.57 | 1.69 | 1.54 | 1.42 | 1.48 | 1.53 | 1.47 | 1.36 |
|  | [1.54-1.60] | [1.66-1.73] | [1.50-1.58] | [1.38-1.46] | [1.44-1.52] | [1.49-1.57] | [1.43-1.51] | [1.32-1.40] |
| **Adjustments** |  |  |  |  |  |  |  |  |
| Sociodemographic | No | Yes | Yes | Yes | Yes | Yes | Yes | Yes |
| Socioeconomic | No | No | Yes | Yes | Yes | Yes | Yes | Yes |
| Health & biomarkers | No | No | No | Yes | No | No | No | Yes |
| Health-related behaviours | No | No | No | No | Yes | No | No | Yes |
| Social isolation | No | No | No | No | No | Yes | No | Yes |
| Psychological | No | No | No | No | No | No | Yes | Yes |
| **Number of Observations** | 432,063 | 425,577 | 356,098 | 325,908 | 330,593 | 354,161 | 321,175 | 275,476 |
| **Number of Patients** | 413,504 | 407,321 | 342,359 | 313,954 | 319,356 | 340,553 | 309,283 | 266,751 |
| **Number of Failures** | 63,662 | 62,713 | 49,367 | 44,122 | 43,133 | 49,001 | 43,418 | 34,678 |
| **Log Likelihood** | -675,012 | -659,292 | -513,512 | -454,767 | -450,316 | -509,498 | -448,161 | -356,025 |

Adjustment for sociodemographic factors strengthened the associations between SCMD and risk of ACSC admissions (Table 2, Additional file Table S3). With the addition of socioeconomic factors, associations remained positive, with the strongest association observed for those with schizophrenia (HR=2.65, 95% CI: 2.37 - 2.97), in models examining the first admission. The associations were attenuated with the addition of health and biomarker variables. Inclusion of the social isolation variables and the psychological variables did not alter associations. The fully adjusted model (Figure 2) indicated reduced associations by around 51%, 37% and 19% among those with schizophrenia, bipolar disorder and anxiety/depression, respectively compared to the unadjusted model. The pattern of results was generally similar when all admissions were considered, but with less attenuation following inclusion of different covariates (Table 3 and Figure 2, Additional file Table S4).

### 3.3 Risk of hospital admissions associated with different covariates

Having a previous diagnosis of a severe (particularly schizophrenia) mental disorder was one of the strongest predictors of hospital admission for ACSCs (Figure 2). Of the sociodemographic factors examined (model 2), increased age, being male and from an ethnic minority group (apart from the White other category) were associated with increased risk of ACSC admission (Additional file Table S3). People from South Asian and Black groups had the highest risk amongst the ethnicity categories investigated. Those living in a rural area were less likely to experience an ACSC admission compared to those from an urban area. Having no educational qualifications, not being in employment and not being a home-owner were all associated with increased risk of ACSC admissions (model 3). Similarly, there was a gradient in the risk of ACSC admission according to household income, with those earning over £100,000 per year showing the lowest risk of ACSC admission compared to those earning less than £18,000.

One of the strongest associations observed amongst the covariates included was when multimorbidity was added to the model. People with four or more physical comorbidities had a threefold higher risk of ACSC admission compared to those without any comorbidity (model 4). Being overweight or obese (but not underweight) were associated with reduced risk of ACSC admission compared to normal weight in models adjusted for sociodemographic and socioeconomic factors. Higher pulse rate, waist circumference and CRP levels were also all associated with increased risk. Previous and current smokers experienced heightened risk of ACSC admissions compared to people who never smoked and former drinkers also had highest risk compared to individuals who had a drink daily or almost daily (model 5). Higher levels of physical activity were related to reduced risk of ACSC.

People who lived with their spouse or partner had lower risk of ACSC admission compared to those who lived alone or with someone else who was not a partner (model 6). Participants who had less social contact with friends or family and who took part in fewer social or leisure activities also had higher risk of ACSC hospitalisation. Loneliness was not related to risk of ACSC admission, whereas experiencing higher levels of depressive symptoms and insomnia were related to increased risk (model 7). In the model containing all predictors (model 8), a SCMD diagnosis was still associated with elevated risk of ACSC admission and most other factors (with the exception of ethnicity, urban/rural status and social participation) also remained associated. Findings were relatively similar when taking all ACSC admissions into account (Additional file Table S4).

## 4 Discussion

### 4.1 Summary of findings

In our study of UK Biobank participants, we found that a previous diagnosis of schizophrenia (or other psychotic disorder) was one of the strongest predictors of being hospitalised for physical health conditions that are amenable to ambulatory care. Bipolar disorder and anxiety/depression were also strongly associated with admissions for ACSCs. Adjustment for socioeconomic circumstances reduced the associations observed, but they persisted even when accounting for a number of different variables, such as biomarkers, health-related behaviours, social isolation and psychological factors. Including health and biomarker variables attenuated the association between SCMD and ACSC admissions, but the inclusion of social isolation and psychological factors (including loneliness) made little difference. Models which considered all hospital admissions for ACSCs, as opposed to just the first event, displayed similar results, although the associations were more conservative overall.

Our findings are in line with the few studies from other countries that demonstrate elevated risk of ACSC-related hospitalisations amongst people with severe mental disorders.(6) Less research has focused on common mental disorders, with most just looking at depression(15). A prior study found elevated levels of ACSC admissions amongst those with anxiety, but this was among a cohort of veterans with diabetes who were not likely to be representative of the wider population.(31) Our study extends this research by including a wide range of covariates which have not been considered in previous studies and by examining all ACSC admissions across a 13-year period. The finding that socioeconomic variables appear to make a key contribution to risk of ACSC admissions amongst those with SCMD suggests more needs to be done to reduce socioeconomic inequalities experienced by those with mental disorders, and in particular people with schizophrenia and other psychotic disorders.

### 4.2 Strengths and limitations

A key strength of our study was the use of UK Biobank data which enabled the exploration of a range of different variables which may influence ACSC admissions. Administrative data alone often lacks detail on important socioeconomic, psychological and health-risk variables,(6, 13) but UK Biobank enabled the linkage of these variables to administrative health records. The large sample size and broad phenotyping provided by UK Biobank also allowed us to examine more detailed psychiatric diagnoses than has been conducted previously, with most prior research combining schizophrenia and bipolar diagnostic groups and not including a comparison to those with common mental disorders, or focusing on depression in isolation.(6, 15) Another significant strength of our analysis was the investigation of multiple hospital admissions per person over a long 13-year follow-up time.

However, a key limitation is that UK Biobank is not representative of the general population; with white, more advantaged and healthier people more likely to participate.(32) This is potentially important for individuals with severe mental illness, as those with more serious illness, who may also have more physical health issues, may be less likely to participate. The consequence of this may be that the associations observed are more likely to be under-, rather than over-estimated. UK Biobank is also susceptible to survival bias as most people were aged 40-70 years at recruitment and we know from previous research that those with severe mental illness are more likely to die prematurely.(2) We excluded those with missing data which may have introduced additional bias to the associations. We have also not conducted a causal mediation analysis which would be needed to elucidate whether the variables included may be mediators of the relationship between mental disorders and ACSC admissions. We also did not investigate whether there were differences in the findings according to factors such as sex and ethnicity which could moderate the association between mental disorders and ACSC admissions, although our models do control for these covariates. Furthermore, we have not explored the specific ACSCs that were associated with a higher burden of hospital admissions; doing so may help to target prevention efforts. Previous research based in Denmark has demonstrated that people with SMI have elevated risk of hospitalisations for acute, infectious illnesses, including appendicitis with perforation, pneumonia, and urinary tract infections.(6)

## 5 Conclusions

People with severe mental disorders had the highest risk of preventable hospital admissions, with the risk also elevated amongst individuals with depression and anxiety. Having a severe mental disorder was also one of the strongest predictors of being admitted to hospital due to an ACSC, despite the inclusion of a wide range of factors. Ensuring those with mental disorders (particularly SMI) receive adequate ambulatory care is essential to reduce the large and growing health inequalities and premature mortality experienced by these groups. Reducing health inequalities among people with SCMD could be achieved by targeting interventions at multiple levels including the individual (e.g. smoking cessation), health system (improved care coordination) and broader society (reduced unemployment, poverty and discrimination).(33) This is particularly pertinent as we emerge from the COVID-19 pandemic, in which there is already substantial evidence that people with mental disorders have been disproportionately affected, in terms of disruptions to healthcare,(34) lower COVID-19 vaccination rates,(35) and adverse outcomes including all-cause and COVID-19 related mortality and hospitalisation.(36, 37)

## Data Availability

The data that support the findings of this study are available from UK Biobank (https://www.ukbiobank.ac.uk/), but restrictions apply to their availability. These data were used under licence for the current study and so are not publicly available. The data are available from the authors upon reasonable request and with permission of UK Biobank.

## 6 List of Abbreviations

ACSCs: Ambulatory Care Sensitive Conditions
BMI: Body Mass Index
CI: Confidence interval
CIPS: Continuous Inpatient Spells
CMD: Common mental disorders
COVID-19: Coronavirus disease-19
CRP: C-Reactive Protein
CSE: Certificate of Secondary Education
GCSE: General Certificate of Secondary Education
GP: General Practice
HES: Hospital Episode Statistics
HR: Hazard Ratio
ICD-10: International Classification of Diseases-10
M: Model
MH: Mental Health
NHS: National Health Service
PH: Proportional Hazard
PHQ: Patient Health Questionnaire
PWP-TT: Prentice, Williams and Peterson Total Time
SMI: Severe mental illness
SCMD: Severe and common mental disorders

## 7 Declarations

### 7.1 Ethics approval and consent to participate

UK Biobank received ethical approval from the NHS National Research Ethics Service North West (21/NW/0157). All participants provided written informed consent before enrolment in the study, which was conducted in accordance with the Declaration of Helsinki.

### 7.2 Consent for publication

Not applicable.

### 7.4 Competing interests

None.

### 7.5 Authors’ contributions

CLN and RJ conceived the idea for the paper. MJA conducted the analysis with assistance from CLN. All authors contributed to the interpretation of the findings. CLN wrote the first draft of the manuscript with assistance from MJA. All authors critically revised the paper for intellectual content. The corresponding author had full access to all the data in the study and had final responsibility for the decision to submit for publication. All authors read and approved the final manuscript and were involved in funding acquisition.

## 7.6 Acknowledgements

We wish to thank the UK Biobank participants. This research has been conducted using the UK Biobank resource under Application 41686 (PI: Niedzwiedz).

## 7.7 Funding

This project has been funded by the Closing the Gap network. Closing the Gap is funded by UK Research and Innovation and their support is gratefully acknowledged (Grant reference: ES/S004459/1). Any views expressed here are those of the project investigators and do not necessarily represent the views of the Closing the Gap network or UKRI. CLN also acknowledges funding from a Medical Research Council Fellowship (MR/R024774/1) and Lord Kelvin/Adam Smith Fellowship.

## Notes

### Competing Interest Statement

Grant funding for research but no other competing interest.

