## Additional file for "Severe and common mental disorders and risk of hospital admissions for Ambulatory Care Sensitive Conditions (ACSCs): prospective cohort study using UK Biobank"

[Table S4: Full results from the Prentice, Williams and Peterson Total Time (PWP-TT] models (all admissions) for the association between SCMD and ACSC admissions	8](#_Toc92115108)

Table S1: HES-APC data: Identify ACSC CIPS

|  | Number of Episodes / CIPS | Number of Patients | SCMD Episodes / CIPS | SCMD Patients |
| --- | --- | --- | --- | --- |
| HES Episodes | 3,341,300 | 392,049 | 952,194 | 87,862 |
| Excluded: |  |  |  |  |
| Unfinished episodes | 409 | 3 | 213 | 0 |
| No start date | 213 | 7 | 65 | 1 |
| No end date | 20 | 0 | 11 | 0 |
| Negative duration | 38 | 0 | 11 | 0 |
| Patients with MH, but non-SMCD diagnosis | 351,044 | 29,866 | 0 | 0 |
| Admission before patient joined biobank | 924,753 | 44,124 | 305,699 | 8,452 |
| After 31 December 2019 | 236,425 | 10,428 | 67,401 | 1,810 |
| Admission before SMI diagnosis | 52,872 | 769 | 52,872 | 769 |
| Elective and maternity | 1,279,270 | 159,818 | 347,066 | 34,546 |
| Duplicates | 445 | 0 | 164 | 0 |
| Final Number of Episodes | 495,811 | 147,034 | 178,692 | 42,284 |
| CIPS | 330,932 | 147,034 | 116,691 | 42,284 |
| ACSC CIPS | 50,677 | 31,787 | 20,651 | 10,859 |

The table above starts with the total number of episodes and patients in HES, then it shows the patients that were excluded due to different reasons. The last three rows show the final number of episodes, after all exclusions, the number of CIPS these episodes form and how many of them are ACSC CIPS.

*Table S2: Number of ACSC admissions among participants people with a severe or common mental disorder (SCMD) diagnosis*

| **Number of Admissions** | **Total Participants** | **Participants with no SCMD** | **Participants with SCMD** |
| --- | --- | --- | --- |
| 0 | 382,220 | 298,526 | 83,694 |
| 1 | 23,192 | 15,974 | 7,218 |
| 2 | 4,769 | 3,010 | 1,759 |
| 3 | 1,691 | 913 | 778 |
| 4 | 818 | 447 | 371 |
| 5 | 416 | 214 | 202 |
| >5 | 785 | 281 | 504 |
| **Total** | **413,891** | **319,365** | **94,526** |

Table S3: Full results from the Cox Proportional Hazard models (first admission) for the association between SCMD and ACSC admissions

|  | **M1** |  |  | **M2** |  |  | **M3** |  |  | **M4** |  |  | **M5** |  |  | **M6** |  |  | **M7** |  |  | **M8** |  |  |
| --- | --- | --- | --- | --- | --- | --- | --- | --- | --- | --- | --- | --- | --- | --- | --- | --- | --- | --- | --- | --- | --- | --- | --- | --- |
|  | **HR** | **L** | **U** | **HR** | **L** | **U** | **HR** | **L** | **U** | **HR** | **L** | **U** | **HR** | **L** | **U** | **HR** | **L** | **U** | **HR** | **L** | **U** | **HR** | **L** | **U** |
| **No SCMD** | 1.00 |  |  | 1.00 |  |  | 1.00 |  |  | 1.00 |  |  | 1.00 |  |  | 1.00 |  |  | 1.00 |  |  | 1.00 |  |  |
| Schizophrenia | 4.40 | 4.04 | 4.80 | 4.86 | 4.44 | 5.32 | 2.65 | 2.37 | 2.97 | 2.31 | 2.05 | 2.61 | 2.47 | 2.19 | 2.79 | 2.65 | 2.36 | 2.97 | 2.56 | 2.26 | 2.89 | 2.15 | 1.87 | 2.48 |
| Bipolar disorder | 2.48 | 2.28 | 2.69 | 2.81 | 2.58 | 3.06 | 2.12 | 1.92 | 2.33 | 1.72 | 1.55 | 1.90 | 1.95 | 1.76 | 2.16 | 2.10 | 1.90 | 2.31 | 1.95 | 1.76 | 2.16 | 1.56 | 1.39 | 1.76 |
| Anxiety or depression | 1.76 | 1.73 | 1.80 | 1.98 | 1.93 | 2.02 | 1.72 | 1.68 | 1.77 | 1.53 | 1.49 | 1.57 | 1.63 | 1.59 | 1.67 | 1.71 | 1.67 | 1.75 | 1.59 | 1.55 | 1.64 | 1.43 | 1.39 | 1.47 |
| Age (continuous) |  |  |  | 1.07 | 1.07 | 1.07 | 1.06 | 1.06 | 1.06 | 1.05 | 1.05 | 1.05 | 1.06 | 1.06 | 1.06 | 1.06 | 1.06 | 1.06 | 1.06 | 1.06 | 1.06 | 1.05 | 1.05 | 1.05 |
| **Sex = Female** |  |  |  | 1.00 |  |  | 1.00 |  |  | 1.00 |  |  | 1.00 |  |  | 1.00 |  |  | 1.00 |  |  | 1.00 |  |  |
| Sex = Male |  |  |  | 1.40 | 1.37 | 1.42 | 1.39 | 1.36 | 1.42 | 1.30 | 1.26 | 1.34 | 1.44 | 1.41 | 1.47 | 1.39 | 1.36 | 1.43 | 1.41 | 1.38 | 1.45 | 1.34 | 1.30 | 1.38 |
| **Ethnicity = White British** |  |  |  | 1.00 |  |  | 1.00 |  |  | 1.00 |  |  | 1.00 |  |  | 1.00 |  |  | 1.00 |  |  | 1.00 |  |  |
| Ethnicity = White Irish |  |  |  | 1.11 | 1.05 | 1.18 | 1.05 | 0.98 | 1.12 | 1.05 | 0.98 | 1.12 | 1.02 | 0.96 | 1.10 | 1.05 | 0.98 | 1.12 | 1.06 | 0.99 | 1.13 | 1.03 | 0.96 | 1.11 |
| Ethnicity = White Other |  |  |  | 0.93 | 0.88 | 0.99 | 0.95 | 0.89 | 1.01 | 0.96 | 0.89 | 1.03 | 0.93 | 0.86 | 0.99 | 0.94 | 0.88 | 1.01 | 0.93 | 0.86 | 0.99 | 0.93 | 0.86 | 1.00 |
| Ethnicity = Mixed |  |  |  | 1.14 | 1.00 | 1.31 | 1.02 | 0.87 | 1.19 | 1.06 | 0.91 | 1.25 | 0.99 | 0.84 | 1.16 | 1.02 | 0.87 | 1.19 | 1.04 | 0.88 | 1.22 | 1.06 | 0.89 | 1.26 |
| Ethnicity = South Asian |  |  |  | 1.39 | 1.30 | 1.48 | 1.18 | 1.09 | 1.28 | 1.09 | 1.00 | 1.19 | 1.07 | 0.98 | 1.17 | 1.19 | 1.10 | 1.29 | 1.10 | 1.00 | 1.21 | 1.02 | 0.91 | 1.13 |
| Ethnicity = Black |  |  |  | 1.38 | 1.28 | 1.49 | 1.07 | 0.97 | 1.17 | 1.07 | 0.97 | 1.18 | 1.06 | 0.96 | 1.16 | 1.08 | 0.98 | 1.18 | 1.07 | 0.97 | 1.19 | 1.07 | 0.96 | 1.20 |
| Ethnicity = Other |  |  |  | 1.20 | 1.10 | 1.31 | 1.03 | 0.92 | 1.15 | 1.05 | 0.93 | 1.18 | 0.96 | 0.85 | 1.07 | 1.04 | 0.93 | 1.16 | 0.98 | 0.87 | 1.11 | 0.95 | 0.83 | 1.10 |
| **Rural = Urban** |  |  |  | 1.00 |  |  | 1.00 |  |  | 1.00 |  |  | 1.00 |  |  | 1.00 |  |  | 1.00 |  |  | 1.00 |  |  |
| Rural = Rural |  |  |  | 0.84 | 0.82 | 0.87 | 0.95 | 0.92 | 0.98 | 0.96 | 0.93 | 0.99 | 0.96 | 0.93 | 0.99 | 0.95 | 0.92 | 0.98 | 0.96 | 0.93 | 0.99 | 0.98 | 0.94 | 1.01 |
| **Education = College or University** |  |  |  |  |  |  | 1.00 |  |  | 1.00 |  |  | 1.00 |  |  | 1.00 |  |  | 1.00 |  |  | 1.00 |  |  |
| Education = A/AS levels |  |  |  |  |  |  | 1.04 | 1.00 | 1.08 | 1.00 | 0.96 | 1.05 | 1.03 | 0.98 | 1.07 | 1.04 | 1.00 | 1.08 | 1.04 | 0.99 | 1.08 | 0.99 | 0.95 | 1.04 |
| Education = O levels / GCSE |  |  |  |  |  |  | 1.11 | 1.08 | 1.15 | 1.07 | 1.03 | 1.10 | 1.09 | 1.06 | 1.12 | 1.11 | 1.08 | 1.15 | 1.10 | 1.06 | 1.13 | 1.04 | 1.00 | 1.08 |
| Education = Other |  |  |  |  |  |  | 1.17 | 1.13 | 1.21 | 1.09 | 1.05 | 1.14 | 1.13 | 1.09 | 1.17 | 1.17 | 1.12 | 1.21 | 1.16 | 1.12 | 1.20 | 1.07 | 1.03 | 1.12 |
| Education = None of the above |  |  |  |  |  |  | 1.26 | 1.22 | 1.30 | 1.17 | 1.13 | 1.21 | 1.19 | 1.15 | 1.23 | 1.25 | 1.21 | 1.29 | 1.23 | 1.18 | 1.27 | 1.12 | 1.07 | 1.16 |
| Townsend deprivation index (continuous) |  |  |  |  |  |  | 1.03 | 1.02 | 1.03 | 1.01 | 1.01 | 1.02 | 1.02 | 1.02 | 1.03 | 1.03 | 1.02 | 1.03 | 1.02 | 1.02 | 1.03 | 1.01 | 1.00 | 1.01 |
| **Employment Status = Paid employment** |  |  |  |  |  |  | 1.00 |  |  | 1.00 |  |  | 1.00 |  |  | 1.00 |  |  | 1.00 |  |  | 1.00 |  |  |
| Employment Status = Retired |  |  |  |  |  |  | 1.13 | 1.10 | 1.17 | 1.06 | 1.03 | 1.09 | 1.13 | 1.09 | 1.16 | 1.15 | 1.12 | 1.18 | 1.12 | 1.08 | 1.15 | 1.05 | 1.02 | 1.09 |
| Employment Status = Looking after home/family |  |  |  |  |  |  | 1.07 | 0.99 | 1.17 | 1.01 | 0.92 | 1.10 | 1.05 | 0.96 | 1.14 | 1.08 | 1.00 | 1.18 | 1.04 | 0.95 | 1.14 | 0.98 | 0.89 | 1.09 |
| Employment Status = Unable to work |  |  |  |  |  |  | 2.24 | 2.13 | 2.35 | 1.52 | 1.45 | 1.61 | 1.94 | 1.83 | 2.05 | 2.24 | 2.13 | 2.36 | 1.97 | 1.86 | 2.08 | 1.39 | 1.31 | 1.49 |
| Employment Status = Unemployed |  |  |  |  |  |  | 1.10 | 1.01 | 1.20 | 1.04 | 0.95 | 1.14 | 1.08 | 0.99 | 1.19 | 1.11 | 1.01 | 1.21 | 1.08 | 0.98 | 1.18 | 1.00 | 0.90 | 1.12 |
| Employment Status = Other |  |  |  |  |  |  | 1.09 | 0.98 | 1.21 | 1.06 | 0.95 | 1.19 | 1.10 | 0.98 | 1.22 | 1.10 | 0.99 | 1.22 | 1.04 | 0.93 | 1.17 | 1.02 | 0.90 | 1.16 |
| **Housing = Own** |  |  |  |  |  |  | 1.00 |  |  | 1.00 |  |  | 1.00 |  |  | 1.00 |  |  | 1.00 |  |  | 1.00 |  |  |
| Housing = Rent/Other |  |  |  |  |  |  | 1.31 | 1.26 | 1.35 | 1.19 | 1.14 | 1.23 | 1.20 | 1.16 | 1.25 | 1.29 | 1.24 | 1.33 | 1.27 | 1.23 | 1.32 | 1.09 | 1.05 | 1.14 |
| **Household Income: < £18,000** |  |  |  |  |  |  | 1.00 |  |  | 1.00 |  |  | 1.00 |  |  | 1.00 |  |  | 1.00 |  |  | 1.00 |  |  |
| HH Income: 18,000 – 30,999 |  |  |  |  |  |  | 0.89 | 0.86 | 0.91 | 0.91 | 0.88 | 0.94 | 0.91 | 0.88 | 0.94 | 0.90 | 0.87 | 0.93 | 0.90 | 0.87 | 0.93 | 0.93 | 0.90 | 0.96 |
| HH Income: 31,000 – 51,999 |  |  |  |  |  |  | 0.83 | 0.80 | 0.86 | 0.86 | 0.83 | 0.89 | 0.86 | 0.83 | 0.89 | 0.84 | 0.81 | 0.87 | 0.84 | 0.81 | 0.87 | 0.89 | 0.85 | 0.93 |
| HH Income: 52,000 – 100,000 |  |  |  |  |  |  | 0.76 | 0.73 | 0.79 | 0.80 | 0.76 | 0.83 | 0.80 | 0.76 | 0.83 | 0.77 | 0.74 | 0.81 | 0.77 | 0.73 | 0.80 | 0.83 | 0.79 | 0.88 |
| HH Income: > 100,000 |  |  |  |  |  |  | 0.71 | 0.66 | 0.76 | 0.76 | 0.70 | 0.81 | 0.76 | 0.71 | 0.82 | 0.73 | 0.68 | 0.78 | 0.73 | 0.68 | 0.78 | 0.80 | 0.74 | 0.86 |
| **Number of Comorbidities = 0** |  |  |  |  |  |  |  |  |  | 1.00 |  |  |  |  |  |  |  |  |  |  |  | 1.00 |  |  |
| Number of Comorbidities = 1 |  |  |  |  |  |  |  |  |  | 1.38 | 1.34 | 1.43 |  |  |  |  |  |  |  |  |  | 1.36 | 1.32 | 1.41 |
| Number of Comorbidities = 2 |  |  |  |  |  |  |  |  |  | 1.76 | 1.70 | 1.82 |  |  |  |  |  |  |  |  |  | 1.71 | 1.65 | 1.78 |
| Number of Comorbidities = 3 |  |  |  |  |  |  |  |  |  | 2.33 | 2.24 | 2.43 |  |  |  |  |  |  |  |  |  | 2.23 | 2.13 | 2.33 |
| Number of Comorbidities = 4+ |  |  |  |  |  |  |  |  |  | 3.15 | 3.01 | 3.29 |  |  |  |  |  |  |  |  |  | 2.89 | 2.75 | 3.05 |
| BMI = Underweight |  |  |  |  |  |  |  |  |  | 2.19 | 1.90 | 2.53 |  |  |  |  |  |  |  |  |  | 2.02 | 1.72 | 2.38 |
| **BMI = Normal** |  |  |  |  |  |  |  |  |  | 1.00 |  |  |  |  |  |  |  |  |  |  |  | 1.00 |  |  |
| BMI = Overweight |  |  |  |  |  |  |  |  |  | 0.84 | 0.81 | 0.87 |  |  |  |  |  |  |  |  |  | 0.86 | 0.83 | 0.90 |
| BMI = Obese |  |  |  |  |  |  |  |  |  | 0.80 | 0.76 | 0.84 |  |  |  |  |  |  |  |  |  | 0.84 | 0.80 | 0.89 |
| Pulse (continuous) |  |  |  |  |  |  |  |  |  | 1.01 | 1.01 | 1.01 |  |  |  |  |  |  |  |  |  | 1.01 | 1.01 | 1.01 |
| Waist circumference (continuous) |  |  |  |  |  |  |  |  |  | 1.01 | 1.01 | 1.01 |  |  |  |  |  |  |  |  |  | 1.01 | 1.01 | 1.01 |
| C-reactive Protein [log] |  |  |  |  |  |  |  |  |  | 1.16 | 1.15 | 1.18 |  |  |  |  |  |  |  |  |  | 1.15 | 1.13 | 1.16 |
| **Smoking = Never** |  |  |  |  |  |  |  |  |  |  |  |  | 1.00 |  |  |  |  |  |  |  |  | 1.00 |  |  |
| Smoking = Previous |  |  |  |  |  |  |  |  |  |  |  |  | 1.24 | 1.21 | 1.27 |  |  |  |  |  |  | 1.15 | 1.12 | 1.18 |
| Smoking = Current |  |  |  |  |  |  |  |  |  |  |  |  | 1.53 | 1.47 | 1.59 |  |  |  |  |  |  | 1.47 | 1.41 | 1.54 |
| **Alcohol = Daily or almost daily** |  |  |  |  |  |  |  |  |  |  |  |  | 1.00 |  |  |  |  |  |  |  |  | 1.00 |  |  |
| Alcohol = 3-4 times/week |  |  |  |  |  |  |  |  |  |  |  |  | 0.95 | 0.91 | 0.98 |  |  |  |  |  |  | 0.96 | 0.92 | 1.00 |
| Alcohol = 1-2 times/week |  |  |  |  |  |  |  |  |  |  |  |  | 1.05 | 1.02 | 1.09 |  |  |  |  |  |  | 1.03 | 0.99 | 1.07 |
| Alcohol = 1-3 times/month |  |  |  |  |  |  |  |  |  |  |  |  | 1.12 | 1.08 | 1.17 |  |  |  |  |  |  | 1.06 | 1.01 | 1.11 |
| Alcohol = Special occasions only |  |  |  |  |  |  |  |  |  |  |  |  | 1.35 | 1.30 | 1.41 |  |  |  |  |  |  | 1.21 | 1.15 | 1.26 |
| Alcohol = Never (former drinker) |  |  |  |  |  |  |  |  |  |  |  |  | 1.51 | 1.43 | 1.59 |  |  |  |  |  |  | 1.34 | 1.26 | 1.42 |
| Alcohol = Never |  |  |  |  |  |  |  |  |  |  |  |  | 1.41 | 1.33 | 1.50 |  |  |  |  |  |  | 1.30 | 1.22 | 1.39 |
| **Physical Activity = 1** |  |  |  |  |  |  |  |  |  |  |  |  | 1.00 |  |  |  |  |  |  |  |  | 1.00 |  |  |
| Physical Activity = 2 |  |  |  |  |  |  |  |  |  |  |  |  | 0.90 | 0.87 | 0.93 |  |  |  |  |  |  | 0.96 | 0.93 | 1.00 |
| Physical Activity = 3 |  |  |  |  |  |  |  |  |  |  |  |  | 0.82 | 0.79 | 0.84 |  |  |  |  |  |  | 0.91 | 0.88 | 0.95 |
| Physical Activity = 4 |  |  |  |  |  |  |  |  |  |  |  |  | 0.82 | 0.79 | 0.85 |  |  |  |  |  |  | 0.95 | 0.92 | 0.99 |
| Physical Activity = 5 |  |  |  |  |  |  |  |  |  |  |  |  | 0.82 | 0.79 | 0.85 |  |  |  |  |  |  | 0.97 | 0.94 | 1.01 |
| **Household Structure = Living with spouse/partner** |  |  |  |  |  |  |  |  |  |  |  |  |  |  |  | 1.00 |  |  |  |  |  | 1.00 |  |  |
| Household Structure = Live with other person |  |  |  |  |  |  |  |  |  |  |  |  |  |  |  | 1.07 | 1.02 | 1.11 |  |  |  | 1.04 | 0.99 | 1.10 |
| Household Structure = Live alone |  |  |  |  |  |  |  |  |  |  |  |  |  |  |  | 1.04 | 1.01 | 1.07 |  |  |  | 1.04 | 1.01 | 1.08 |
| **Visits friends/family ≥ 1/week** |  |  |  |  |  |  |  |  |  |  |  |  |  |  |  | 1.00 |  |  |  |  |  | 1.00 |  |  |
| Visits friends/family < 1/week |  |  |  |  |  |  |  |  |  |  |  |  |  |  |  | 1.03 | 1.01 | 1.06 |  |  |  | 1.04 | 1.01 | 1.07 |
| **Leisure/social activities ≥ 1/week** |  |  |  |  |  |  |  |  |  |  |  |  |  |  |  | 1.00 |  |  |  |  |  | 1.00 |  |  |
| Leisure/social activities < 1/week |  |  |  |  |  |  |  |  |  |  |  |  |  |  |  | 1.13 | 1.10 | 1.16 |  |  |  | 0.99 | 0.97 | 1.02 |
| **Lonely = No** |  |  |  |  |  |  |  |  |  |  |  |  |  |  |  |  |  |  | 1.00 |  |  | 1.00 |  |  |
| Lonely = Yes |  |  |  |  |  |  |  |  |  |  |  |  |  |  |  |  |  |  | 1.00 | 0.97 | 1.03 | 0.99 | 0.95 | 1.02 |
| PHQ (continuous) |  |  |  |  |  |  |  |  |  |  |  |  |  |  |  |  |  |  | 1.05 | 1.05 | 1.06 | 1.02 | 1.01 | 1.03 |
| **Insomnia = Never/rarely** |  |  |  |  |  |  |  |  |  |  |  |  |  |  |  |  |  |  | 1.00 |  |  | 1.00 |  |  |
| Insomnia = Sometimes |  |  |  |  |  |  |  |  |  |  |  |  |  |  |  |  |  |  | 1.01 | 0.98 | 1.04 | 1.00 | 0.97 | 1.03 |
| Insomnia = Usually |  |  |  |  |  |  |  |  |  |  |  |  |  |  |  |  |  |  | 1.11 | 1.07 | 1.14 | 1.04 | 1.01 | 1.08 |

ACSC=Ambulatory Care Sensitive Conditions; BMI=Body Mass Index; HH=Household; HR=Hazard ratio; L=Lower 95% confidence interval; M=Model; PHQ=Patient Health Questionnaire; SCMD=Severe and common mental disorders; U=Upper 95% confidence interval; Reference categories are highlighted in bold

Table S4: Full results from the Prentice, Williams and Peterson Total Time (PWP-TT] models (all admissions) for the association between SCMD and ACSC admissions

|  | **M1** |  |  | **M2** |  |  | **M3** |  |  | **M4** |  |  | **M5** |  |  | **M6** |  |  | **M7** |  |  | **M8** |  |  |
| --- | --- | --- | --- | --- | --- | --- | --- | --- | --- | --- | --- | --- | --- | --- | --- | --- | --- | --- | --- | --- | --- | --- | --- | --- |
|  | **HR** | **L** | **U** | **HR** | **L** | **U** | **HR** | **L** | **U** | **HR** | **L** | **U** | **HR** | **L** | **U** | **HR** | **L** | **U** | **HR** | **L** | **U** | **HR** | **L** | **U** |
| **No SCMD** | 1.00 |  |  | 1.00 |  |  | 1.00 |  |  | 1.00 |  |  | 1.00 |  |  | 1.00 |  |  | 1.00 |  |  | 1.00 |  |  |
| Schizophrenia | 2.29 | 2.08 | 2.52 | 2.64 | 2.38 | 2.93 | 2.09 | 1.84 | 2.38 | 1.96 | 1.70 | 2.25 | 1.92 | 1.66 | 2.23 | 2.10 | 1.85 | 2.39 | 2.11 | 1.83 | 2.42 | 1.86 | 1.57 | 2.20 |
| Bipolar disorder | 1.92 | 1.77 | 2.08 | 2.13 | 1.95 | 2.33 | 1.79 | 1.62 | 1.98 | 1.58 | 1.42 | 1.76 | 1.84 | 1.68 | 2.03 | 1.78 | 1.61 | 1.97 | 1.73 | 1.55 | 1.92 | 1.62 | 1.46 | 1.80 |
| Anxiety or depression | 1.57 | 1.54 | 1.60 | 1.69 | 1.66 | 1.73 | 1.54 | 1.50 | 1.58 | 1.42 | 1.38 | 1.46 | 1.48 | 1.44 | 1.52 | 1.53 | 1.49 | 1.57 | 1.47 | 1.43 | 1.51 | 1.36 | 1.32 | 1.40 |
| Age (continuous) |  |  |  | 1.05 | 1.05 | 1.05 | 1.05 | 1.04 | 1.05 | 1.04 | 1.04 | 1.04 | 1.05 | 1.05 | 1.05 | 1.05 | 1.05 | 1.05 | 1.05 | 1.05 | 1.05 | 1.04 | 1.04 | 1.05 |
| **Sex = Female** |  |  |  | 1.00 |  |  | 1.00 |  |  | 1.00 |  |  | 1.00 |  |  | 1.00 |  |  | 1.00 |  |  | 1.00 |  |  |
| Sex = Male |  |  |  | 1.30 | 1.27 | 1.32 | 1.29 | 1.27 | 1.32 | 1.25 | 1.21 | 1.29 | 1.34 | 1.31 | 1.37 | 1.30 | 1.27 | 1.33 | 1.31 | 1.28 | 1.35 | 1.27 | 1.23 | 1.32 |
| **Ethnicity = White British** |  |  |  | 1.00 |  |  | 1.00 |  |  | 1.00 |  |  | 1.00 |  |  | 1.00 |  |  | 1.00 |  |  | 1.00 |  |  |
| Ethnicity = White Irish |  |  |  | 1.08 | 1.01 | 1.15 | 1.03 | 0.96 | 1.11 | 1.01 | 0.93 | 1.10 | 1.02 | 0.94 | 1.10 | 1.03 | 0.96 | 1.11 | 1.02 | 0.95 | 1.11 | 1.00 | 0.91 | 1.09 |
| Ethnicity = White Other |  |  |  | 0.97 | 0.92 | 1.03 | 1.00 | 0.94 | 1.07 | 0.99 | 0.92 | 1.07 | 0.99 | 0.93 | 1.07 | 1.00 | 0.93 | 1.06 | 0.97 | 0.91 | 1.05 | 0.99 | 0.91 | 1.07 |
| Ethnicity = Mixed |  |  |  | 1.06 | 0.93 | 1.22 | 1.00 | 0.85 | 1.17 | 1.04 | 0.91 | 1.19 | 1.03 | 0.89 | 1.18 | 1.01 | 0.86 | 1.18 | 1.07 | 0.91 | 1.25 | 1.07 | 0.93 | 1.24 |
| Ethnicity = South Asian |  |  |  | 1.16 | 1.08 | 1.25 | 1.03 | 0.94 | 1.13 | 0.99 | 0.90 | 1.08 | 0.97 | 0.88 | 1.07 | 1.03 | 0.94 | 1.13 | 0.98 | 0.88 | 1.10 | 0.98 | 0.88 | 1.10 |
| Ethnicity = Black |  |  |  | 1.32 | 1.22 | 1.42 | 1.13 | 1.03 | 1.23 | 1.13 | 1.03 | 1.24 | 1.16 | 1.05 | 1.27 | 1.14 | 1.04 | 1.24 | 1.13 | 1.03 | 1.25 | 1.19 | 1.07 | 1.33 |
| Ethnicity = Other |  |  |  | 1.17 | 1.07 | 1.27 | 1.01 | 0.91 | 1.12 | 1.06 | 0.95 | 1.18 | 0.99 | 0.89 | 1.10 | 1.03 | 0.93 | 1.14 | 0.95 | 0.84 | 1.06 | 0.97 | 0.85 | 1.10 |
| **Rural = Urban** |  |  |  | 1.00 |  |  | 1.00 |  |  | 1.00 |  |  | 1.00 |  |  | 1.00 |  |  | 1.00 |  |  | 1.00 |  |  |
| Rural = Rural |  |  |  | 0.88 | 0.85 | 0.91 | 0.97 | 0.93 | 1.00 | 0.97 | 0.94 | 1.00 | 0.97 | 0.93 | 1.00 | 0.97 | 0.93 | 1.00 | 0.97 | 0.94 | 1.01 | 0.98 | 0.95 | 1.02 |
| **Education = College or University** |  |  |  |  |  |  | 1.00 |  |  | 1.00 |  |  | 1.00 |  |  | 1.00 |  |  | 1.00 |  |  | 1.00 |  |  |
| Education = A/AS levels |  |  |  |  |  |  | 1.06 | 1.02 | 1.10 | 1.03 | 0.98 | 1.07 | 1.05 | 1.01 | 1.09 | 1.06 | 1.02 | 1.10 | 1.06 | 1.01 | 1.10 | 1.01 | 0.97 | 1.06 |
| Education = O levels / GCSE |  |  |  |  |  |  | 1.10 | 1.07 | 1.14 | 1.06 | 1.03 | 1.10 | 1.08 | 1.05 | 1.12 | 1.10 | 1.07 | 1.14 | 1.09 | 1.05 | 1.12 | 1.04 | 1.00 | 1.07 |
| Education = Other |  |  |  |  |  |  | 1.14 | 1.10 | 1.18 | 1.08 | 1.04 | 1.13 | 1.10 | 1.06 | 1.14 | 1.14 | 1.10 | 1.18 | 1.13 | 1.08 | 1.17 | 1.04 | 0.99 | 1.08 |
| Education = None of the above |  |  |  |  |  |  | 1.19 | 1.15 | 1.23 | 1.14 | 1.10 | 1.18 | 1.14 | 1.10 | 1.18 | 1.17 | 1.13 | 1.22 | 1.17 | 1.12 | 1.21 | 1.09 | 1.05 | 1.14 |
| Townsend deprivation index (continuous) |  |  |  |  |  |  | 1.02 | 1.01 | 1.02 | 1.01 | 1.00 | 1.01 | 1.01 | 1.01 | 1.02 | 1.02 | 1.01 | 1.02 | 1.01 | 1.01 | 1.02 | 1.00 | 1.00 | 1.01 |
| **Employment Status = Paid employment** |  |  |  |  |  |  | 1.00 |  |  | 1.00 |  |  | 1.00 |  |  | 1.00 |  |  | 1.00 |  |  | 1.00 |  |  |
| Employment Status = Retired |  |  |  |  |  |  | 1.11 | 1.07 | 1.14 | 1.03 | 1.00 | 1.06 | 1.08 | 1.05 | 1.12 | 1.12 | 1.09 | 1.15 | 1.09 | 1.06 | 1.13 | 1.02 | 0.98 | 1.06 |
| Employment Status = Looking after home/family |  |  |  |  |  |  | 1.10 | 1.02 | 1.20 | 1.05 | 0.97 | 1.15 | 1.08 | 0.99 | 1.18 | 1.12 | 1.03 | 1.21 | 1.07 | 0.98 | 1.17 | 1.04 | 0.94 | 1.15 |
| Employment Status = Unable to work |  |  |  |  |  |  | 1.63 | 1.55 | 1.72 | 1.28 | 1.21 | 1.36 | 1.51 | 1.43 | 1.60 | 1.64 | 1.56 | 1.73 | 1.49 | 1.41 | 1.58 | 1.22 | 1.13 | 1.30 |
| Employment Status = Unemployed |  |  |  |  |  |  | 1.21 | 1.11 | 1.32 | 1.13 | 1.04 | 1.23 | 1.20 | 1.10 | 1.30 | 1.22 | 1.12 | 1.33 | 1.21 | 1.10 | 1.33 | 1.09 | 0.99 | 1.21 |
| Employment Status = Other |  |  |  |  |  |  | 1.19 | 1.07 | 1.32 | 1.12 | 1.00 | 1.26 | 1.18 | 1.05 | 1.33 | 1.20 | 1.08 | 1.33 | 1.15 | 1.02 | 1.29 | 1.06 | 0.92 | 1.22 |
| **Housing = Own** |  |  |  |  |  |  | 1.00 |  |  | 1.00 |  |  | 1.00 |  |  | 1.00 |  |  | 1.00 |  |  | 1.00 |  |  |
| Housing = Rent/Other |  |  |  |  |  |  | 1.15 | 1.10 | 1.19 | 1.07 | 1.03 | 1.11 | 1.09 | 1.05 | 1.13 | 1.13 | 1.08 | 1.17 | 1.13 | 1.08 | 1.18 | 0.99 | 0.94 | 1.03 |
| **Household Income: < £18,000** |  |  |  |  |  |  | 1.00 |  |  | 1.00 |  |  | 1.00 |  |  | 1.00 |  |  | 1.00 |  |  | 1.00 |  |  |
| HH Income: 18,000 – 30,999 |  |  |  |  |  |  | 0.92 | 0.90 | 0.95 | 0.93 | 0.90 | 0.96 | 0.94 | 0.91 | 0.97 | 0.93 | 0.90 | 0.96 | 0.94 | 0.91 | 0.97 | 0.95 | 0.91 | 0.98 |
| HH Income: 31,000 – 51,999 |  |  |  |  |  |  | 0.85 | 0.82 | 0.88 | 0.87 | 0.84 | 0.90 | 0.87 | 0.84 | 0.91 | 0.86 | 0.83 | 0.89 | 0.87 | 0.84 | 0.90 | 0.90 | 0.86 | 0.93 |
| HH Income: 52,000 – 100,000 |  |  |  |  |  |  | 0.76 | 0.73 | 0.79 | 0.79 | 0.75 | 0.83 | 0.80 | 0.76 | 0.83 | 0.78 | 0.74 | 0.81 | 0.77 | 0.74 | 0.81 | 0.83 | 0.79 | 0.87 |
| HH Income: > 100,000 |  |  |  |  |  |  | 0.68 | 0.63 | 0.73 | 0.72 | 0.67 | 0.78 | 0.72 | 0.67 | 0.78 | 0.69 | 0.65 | 0.75 | 0.70 | 0.65 | 0.75 | 0.77 | 0.71 | 0.83 |
| **Number of Comorbidities = 0** |  |  |  |  |  |  |  |  |  | 1.00 |  |  |  |  |  |  |  |  |  |  |  | 1.00 |  |  |
| Number of Comorbidities = 1 |  |  |  |  |  |  |  |  |  | 1.43 | 1.38 | 1.48 |  |  |  |  |  |  |  |  |  | 1.40 | 1.35 | 1.45 |
| Number of Comorbidities = 2 |  |  |  |  |  |  |  |  |  | 1.80 | 1.74 | 1.87 |  |  |  |  |  |  |  |  |  | 1.75 | 1.68 | 1.82 |
| Number of Comorbidities = 3 |  |  |  |  |  |  |  |  |  | 2.17 | 2.08 | 2.27 |  |  |  |  |  |  |  |  |  | 2.06 | 1.97 | 2.16 |
| Number of Comorbidities = 4+ |  |  |  |  |  |  |  |  |  | 2.31 | 2.19 | 2.43 |  |  |  |  |  |  |  |  |  | 2.18 | 2.06 | 2.31 |
| BMI = Underweight |  |  |  |  |  |  |  |  |  | 1.88 | 1.61 | 2.18 |  |  |  |  |  |  |  |  |  | 1.69 | 1.40 | 2.04 |
| **BMI = Normal** |  |  |  |  |  |  |  |  |  | 1.00 |  |  |  |  |  |  |  |  |  |  |  | 1.00 |  |  |
| BMI = Overweight |  |  |  |  |  |  |  |  |  | 0.89 | 0.86 | 0.92 |  |  |  |  |  |  |  |  |  | 0.91 | 0.88 | 0.94 |
| BMI = Obese |  |  |  |  |  |  |  |  |  | 0.86 | 0.82 | 0.90 |  |  |  |  |  |  |  |  |  | 0.90 | 0.85 | 0.95 |
| Pulse (continuous) |  |  |  |  |  |  |  |  |  | 1.01 | 1.00 | 1.01 |  |  |  |  |  |  |  |  |  | 1.01 | 1.00 | 1.01 |
| Waist circumference (continuous) |  |  |  |  |  |  |  |  |  | 1.01 | 1.01 | 1.01 |  |  |  |  |  |  |  |  |  | 1.01 | 1.01 | 1.01 |
| C-reactive Protein [log] |  |  |  |  |  |  |  |  |  | 1.12 | 1.11 | 1.13 |  |  |  |  |  |  |  |  |  | 1.11 | 1.09 | 1.12 |
| **Smoking = Never** |  |  |  |  |  |  |  |  |  |  |  |  | 1.00 |  |  |  |  |  |  |  |  | 1.00 |  |  |
| Smoking = Previous |  |  |  |  |  |  |  |  |  |  |  |  | 1.17 | 1.15 | 1.20 |  |  |  |  |  |  | 1.12 | 1.09 | 1.15 |
| Smoking = Current |  |  |  |  |  |  |  |  |  |  |  |  | 1.39 | 1.34 | 1.45 |  |  |  |  |  |  | 1.40 | 1.34 | 1.47 |
| **Alcohol = Daily or almost daily** |  |  |  |  |  |  |  |  |  |  |  |  | 1.00 |  |  |  |  |  |  |  |  | 1.00 |  |  |
| Alcohol = 3-4 times/week |  |  |  |  |  |  |  |  |  |  |  |  | 0.94 | 0.90 | 0.97 |  |  |  |  |  |  | 0.96 | 0.93 | 1.00 |
| Alcohol = 1-2 times/week |  |  |  |  |  |  |  |  |  |  |  |  | 1.04 | 1.00 | 1.07 |  |  |  |  |  |  | 1.04 | 1.00 | 1.08 |
| Alcohol = 1-3 times/month |  |  |  |  |  |  |  |  |  |  |  |  | 1.08 | 1.03 | 1.13 |  |  |  |  |  |  | 1.05 | 1.01 | 1.11 |
| Alcohol = Special occasions only |  |  |  |  |  |  |  |  |  |  |  |  | 1.24 | 1.19 | 1.29 |  |  |  |  |  |  | 1.14 | 1.09 | 1.20 |
| Alcohol = Never (former drinker) |  |  |  |  |  |  |  |  |  |  |  |  | 1.30 | 1.23 | 1.38 |  |  |  |  |  |  | 1.20 | 1.13 | 1.29 |
| Alcohol = Never |  |  |  |  |  |  |  |  |  |  |  |  | 1.31 | 1.23 | 1.39 |  |  |  |  |  |  | 1.24 | 1.16 | 1.33 |
| **Physical Activity = 1** |  |  |  |  |  |  |  |  |  |  |  |  | 1.00 |  |  |  |  |  |  |  |  | 1.00 |  |  |
| Physical Activity = 2 |  |  |  |  |  |  |  |  |  |  |  |  | 0.93 | 0.90 | 0.96 |  |  |  |  |  |  | 0.98 | 0.94 | 1.02 |
| Physical Activity = 3 |  |  |  |  |  |  |  |  |  |  |  |  | 0.86 | 0.83 | 0.89 |  |  |  |  |  |  | 0.94 | 0.90 | 0.98 |
| Physical Activity = 4 |  |  |  |  |  |  |  |  |  |  |  |  | 0.88 | 0.85 | 0.91 |  |  |  |  |  |  | 0.98 | 0.94 | 1.02 |
| Physical Activity = 5 |  |  |  |  |  |  |  |  |  |  |  |  | 0.89 | 0.86 | 0.93 |  |  |  |  |  |  | 1.02 | 0.98 | 1.06 |
| **Household Structure = Living with spouse/partner** |  |  |  |  |  |  |  |  |  |  |  |  |  |  |  | 1.00 |  |  |  |  |  | 1.00 |  |  |
| Household Structure = Live with other person |  |  |  |  |  |  |  |  |  |  |  |  |  |  |  | 1.08 | 1.03 | 1.12 |  |  |  | 1.06 | 1.01 | 1.12 |
| Household Structure = Live alone |  |  |  |  |  |  |  |  |  |  |  |  |  |  |  | 1.03 | 1.00 | 1.06 |  |  |  | 1.03 | 1.00 | 1.07 |
| **Visits friends/family ≥ 1/week** |  |  |  |  |  |  |  |  |  |  |  |  |  |  |  | 1.00 |  |  |  |  |  | 1.00 |  |  |
| Visits friends/family < 1/week |  |  |  |  |  |  |  |  |  |  |  |  |  |  |  | 1.04 | 1.01 | 1.07 |  |  |  | 1.05 | 1.02 | 1.08 |
| **Leisure/social activities ≥ 1/week** |  |  |  |  |  |  |  |  |  |  |  |  |  |  |  | 1.00 |  |  |  |  |  | 1.00 |  |  |
| Leisure/social activities < 1/week |  |  |  |  |  |  |  |  |  |  |  |  |  |  |  | 1.10 | 1.07 | 1.12 |  |  |  | 1.00 | 0.97 | 1.03 |
| **Lonely = No** |  |  |  |  |  |  |  |  |  |  |  |  |  |  |  |  |  |  | 1.00 |  |  | 1.00 |  |  |
| Lonely = Yes |  |  |  |  |  |  |  |  |  |  |  |  |  |  |  |  |  |  | 1.00 | 0.97 | 1.04 | 0.99 | 0.95 | 1.03 |
| PHQ (continuous) |  |  |  |  |  |  |  |  |  |  |  |  |  |  |  |  |  |  | 1.03 | 1.03 | 1.04 | 1.01 | 1.00 | 1.02 |
| **Insomnia = Never/rarely** |  |  |  |  |  |  |  |  |  |  |  |  |  |  |  |  |  |  | 1.00 |  |  | 1.00 |  |  |
| Insomnia = Sometimes |  |  |  |  |  |  |  |  |  |  |  |  |  |  |  |  |  |  | 1.00 | 0.97 | 1.03 | 0.99 | 0.95 | 1.02 |
| Insomnia = Usually |  |  |  |  |  |  |  |  |  |  |  |  |  |  |  |  |  |  | 1.06 | 1.03 | 1.10 | 1.01 | 0.97 | 1.05 |

ACSC=Ambulatory Care Sensitive conditions; BMI=Body Mass Index; HH=Household; HR=Hazard ratio; L=Lower 95% confidence interval; M=Model; PHQ=Patient Health Questionnaire; SCMD=Severe and common mental disorders; U=Upper 95% confidence interval; Reference categories are highlighted in bold
